# Reproducibility-driven discovery and systematic benchmarking reveal a robust cerebrospinal fluid proteomic signature in Alzheimer’s disease

**DOI:** 10.64898/2026.04.26.26351766

**Authors:** María Fernanda Zambrano-Astorga, Aldo Moreno-Ulloa, the Alzheimer’s Disease Neuroimaging Initiative

## Abstract

Numerous cerebrospinal fluid (CSF) proteomic signatures for Alzheimer’s disease (AD) diagnosis and prognosis have been proposed. However, cross-cohort reproducibility and head-to-head comparison among signatures remain uncertain. We implemented a reproducibility-driven framework integrating systematic review, multi-cohort validation, and systematic benchmarking to prioritized robust biomarkers. Across eight discovery studies (n=759) we identified eleven consistently dysregulated proteins (termed PPAV11). In three independent validation cohorts (n=1,198), PPAV11 demonstrated high diagnostic accuracy (AUC>0.94) and significant prognostic capacity (CU to A⁺T⁺ MCI HR>4.96, *p*=0.004; A⁺T⁺ MCI to A⁺T⁺ dementia HR>3.23, *p*=3.13×10⁻⁷). Comparative benchmarking against thirteen published signatures revealed superior cross-context stability across diagnostic definitions, disease stages, and proteomic platforms. Biologically, PPAV11 captures synaptic, metabolic, immune, and vascular processes and correlates with cognitive decline and neurodegeneration. Together, these findings establish reproducibility as an important criterion for proteomic biomarker prioritization and define a stable molecular signature for integrated AD diagnosis and prognosis.

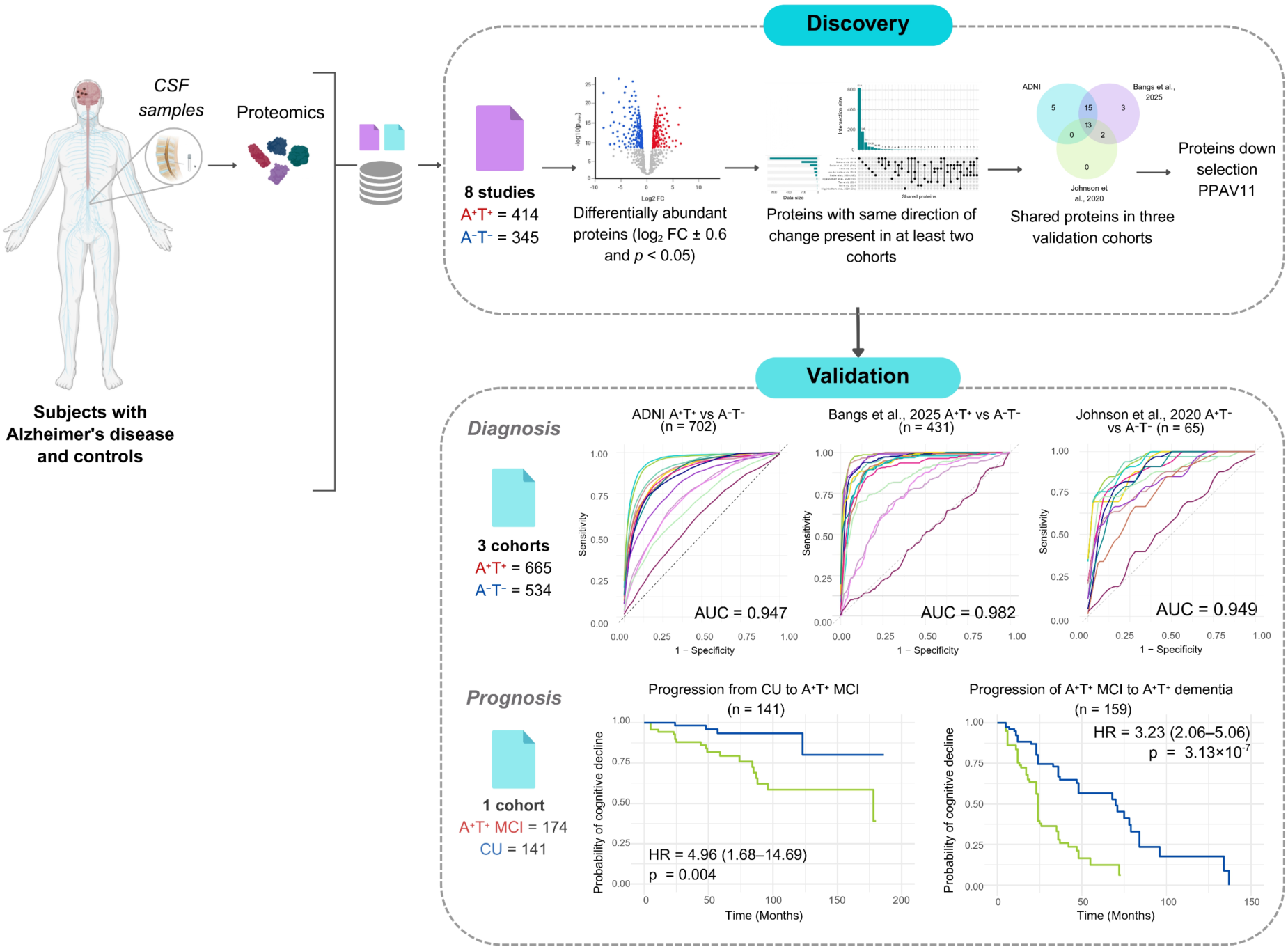

**Highlights:**

1. A reproducibility framework identifies stable CSF proteomic signature for AD.
2. PPAV11 shows strong diagnostic accuracy across cohorts, platforms, and stratifications.
3. PPAV11 levels correlate with cognitive decline and predict AD progression.
4. PPAV11 unites synaptic, metabolic, immune, and vascular pathways in AD dynamics.

## Introduction

Alzheimer’s disease (AD) is neuropathologically characterized by the accumulation of amyloid beta (Aβ) plaques and phosphorylated Tau (p-Tau) tangles in the brain. Recently, the National Institute on Aging and the Alzheimer’s Association (NIA-AA) proposed a biological definition of AD based exclusively on the presence of abnormal Aβ biomarkers, independent of clinical symptoms^1^. In contrast, the International Working Group (IWG) recommends diagnosing AD through the combined assessment of clinical manifestations and the presence of Aβ and Tau pathology^2^. While the NIA-AA framework emphasizes the transition toward biomarker-based diagnosis, the IWG highlights the continued importance of clinical presentation. Although Aβ and p-Tau biomarkers are highly effective for detecting core AD pathology, they explain only 20–40% of the variance in cognitive impairment (CI) and provide limited insight into disease severity and progression^3–5^. This limited explanatory power suggests involvement of additional biological mechanisms driving neurodegeneration and cognitive decline. Indeed, converging evidence indicates that AD arises from a complex network of biochemical and cellular processes—including innate immune activation, lipid metabolism, synaptic dysfunction, RNA regulation, vascular biology, and blood–brain barrier integrity—that extend beyond Aβ and p-Tau dysregulation^4,6,7^. In addition, multi-omics studies have identified numerous genetic risk loci, distinct molecular AD subtypes, and biological modules, supporting the existence of multiple pathogenic cascades that differentially contribute to cognitive decline and neurodegeneration^4,6,8–13^. Despite increasing recognition of AD’s biological heterogeneity, biomarkers that reliably associate molecular alterations with cognitive decline and neurodegeneration remain limited. This disconnect constrains patient stratification, clinical staging, and the assessment of disease progression and therapeutic response, underscoring the need for biomarkers that capture biological processes beyond Aβ and p-Tau pathologies^4,14–16^. Proteomics offers a powerful and unbiased approach for the discovery of such biomarkers. Proteins represent the direct effectors of genetic and environmental risk factors and may therefore accurately reflect disease-relevant biological processes^17^. Fluid-based proteomics is particularly attractive, as it enables the simultaneous assessment of multiple biological pathways from a single sample. Among biological fluids, cerebrospinal fluid (CSF) is especially informative in the study of neurodegenerative diseases^8,18,19^ due to its proximity and physiological connectivity to the brain^20^. Accordingly, numerous CSF proteomic studies have proposed protein signatures for AD diagnosis and prognosis^6,8,12,21–28^. However, these studies are frequently limited by small sample sizes, methodological heterogeneity, and inconsistent diagnostic definitions, which collectively undermine reproducibility and comparability^15,29^. Moreover, although those proteomics signatures have shown good-to-excellent diagnostic and prognostic performance in independent reports, no head-to-head comparisons have evaluated their relative utility—further limiting insights into their true potential clinical value. To address these shortcomings, we conducted this study with two aims: (1) a comprehensive systematic review of publicly available CSF proteomic datasets to identify reproducible protein biomarkers that capture AD’s multifaceted biology and align with its clinical manifestations; and (2) a head-to-head comparison of previously published proteomic signatures to rigorously assess their reproducibility, diagnostic accuracy, and prognostic performance across independent cohorts. Together, this approach seeks to clarify the robustness and clinical relevance of CSF proteomic biomarkers for AD and to prioritize candidates for validation in larger, prospective studies.

## Results

### Study design, systematic review, and data selection

We implemented a four-stage analytical workflow (**Figure 1**). (1) We first conducted a systematic review to identify CSF-based proteomic studies that compared individuals with AD to controls. A total of 484 studies were identified. We applied predefined inclusion and exclusion criteria to ensure methodological consistency. (2) From eight studies that met our criteria (subjects, n = 759) (**Figures 2A and 2B**), we performed a reproducibility-based analysis to identify proteins that were consistently dysregulated across datasets. This involved using combined biological and statistical filtering. Of those subjects, 414 and 345 were diagnosed as A^+^T^+^ and A⁻T⁻, respectively. (3) We then evaluated the diagnostic and prognostic performance of the resulting protein signature across four independent datasets, including three AD cohorts (n = 1,198) and one Parkinson’s disease (PD) cohort (n = 798) for differential diagnosis. For diagnostic performance, subjects were stratified according to biological (A^+^T^+^) and combined biological and clinical (A^+^T^+^ plus cognitive impairment, CI) definitions of AD, using binary classification models and receiver operating characteristic (ROC) curve analyses. Diagnostic specificity was further assessed against PD (n = 798) and in subsets of individuals with CI not linked to AD (A⁻T⁻, n = 488), and prognostic utility was evaluated for predicting cognitive decline implementing the Cox proportional hazards model.

**Figure 1.**
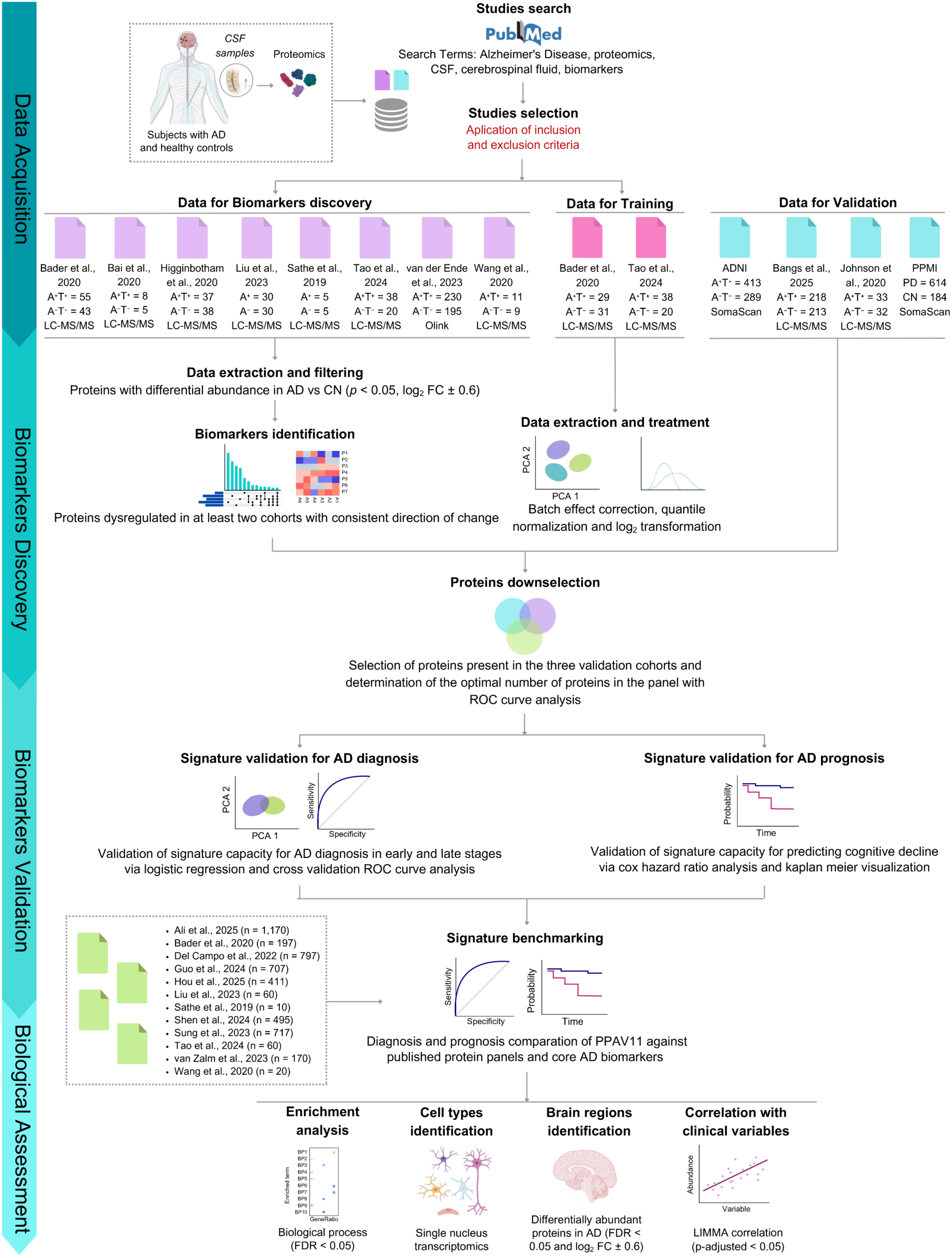
Overview of the experimental and analytical pipeline. A four-step approach was followed, including data selection for analysis, protein signature discovery, validation in independent cohorts and benchmarking against published protein signatures, and evaluation of the biological and clinical relevance of the protein signature (referred to as PPAV11). Abbreviations: AD, Alzheimer’s disease; A, amyloid beta; T, Tau; CN, control; ROC, receiver operating characteristics; ADNI, Alzheimer’s Disease Neuroimaging Initiative; PPMI, Parkinson’s Progression Markers Initiative; LC-MS/MS, liquid chromatography coupled to Tandem mass spectrometry; CI, confidence intervals; CSF, cerebrospinal fluid; FC, fold change; PCA, principal component analysis. Figure was created with biorender.com.

**Figure 2.**
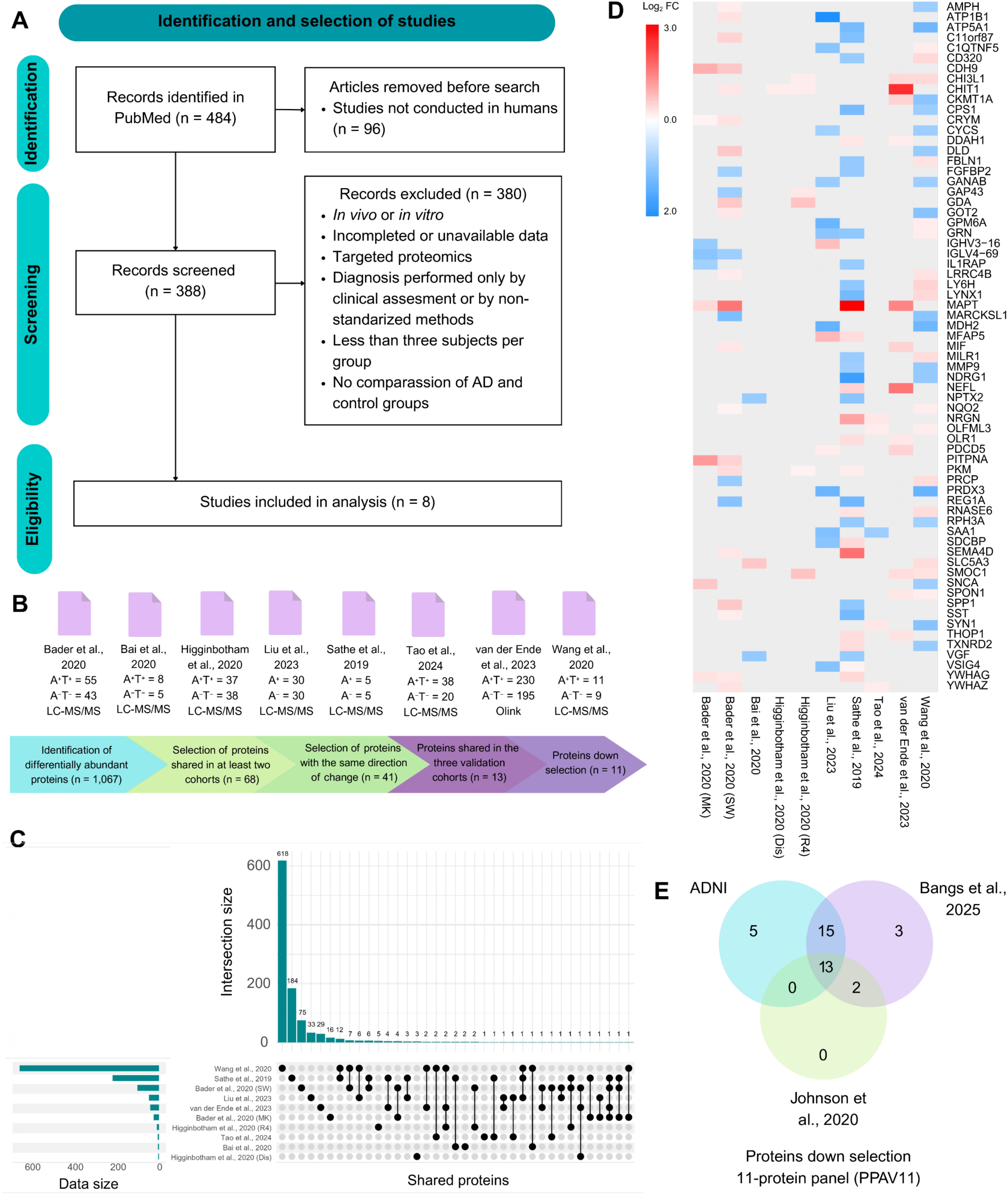
Reproducibility-centered approach for the discovery of a proteomic signature for Alzheimer’s Disease (AD) diagnosis and prognosis. (A) Systematic review of proteomics studies on cerebrospinal fluid using PubMed. (B) Studies meeting eligibility criteria for the discovery step and description of the process to identify proteins composing the PPAV11 signature. (C) Overlap of dysregulated proteins (log^2^ FC ± 0.6 and *p* < 0.05) between AD and control subjects across discovery studies. (D) Heatmap of proteins from (C) present in at least two studies. (E) Overlapping dysregulated proteins with consistent direction of change between AD and control subjects in validation cohorts.

Participants represented a variety of ethnic backgrounds being predominant by Caucasians. Among discovery and validation studies reporting sex distribution, a comparable representation was observed, with 1,227 females and 1,486 males. Detailed sex proportions and subjects numbers for each cohort are described in **Table 1**. The results were compared with thirteen previously published proteomic signatures. (4) Finally, we investigated the molecular, cellular, and physiological insights of the identified proteins. We used gene ontology (GO) and pathway enrichment analyses, single-nucleus brain transcriptomic data, and correlation analyses with clinical variables to enhance our understanding.

**Table 1.**
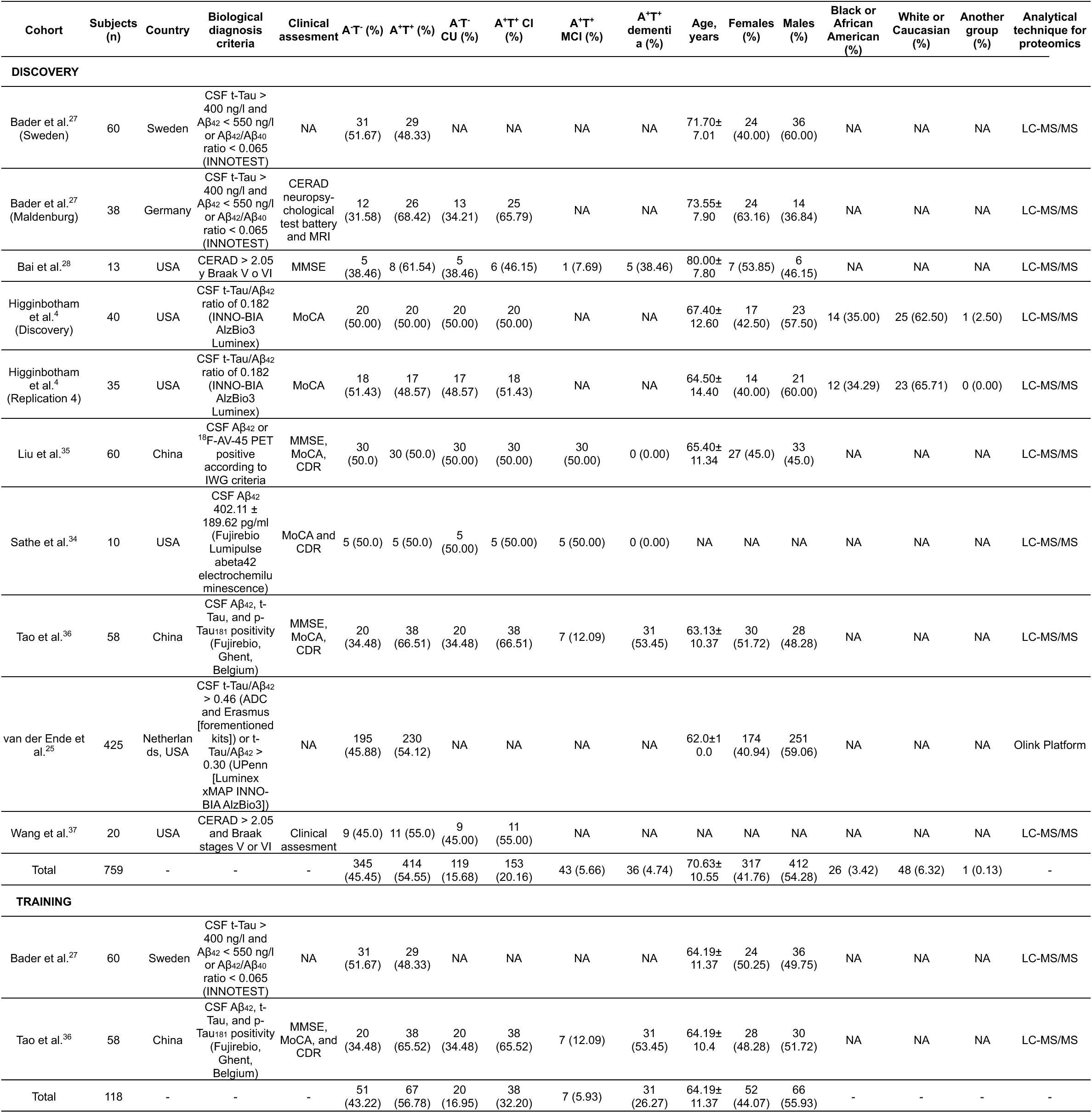
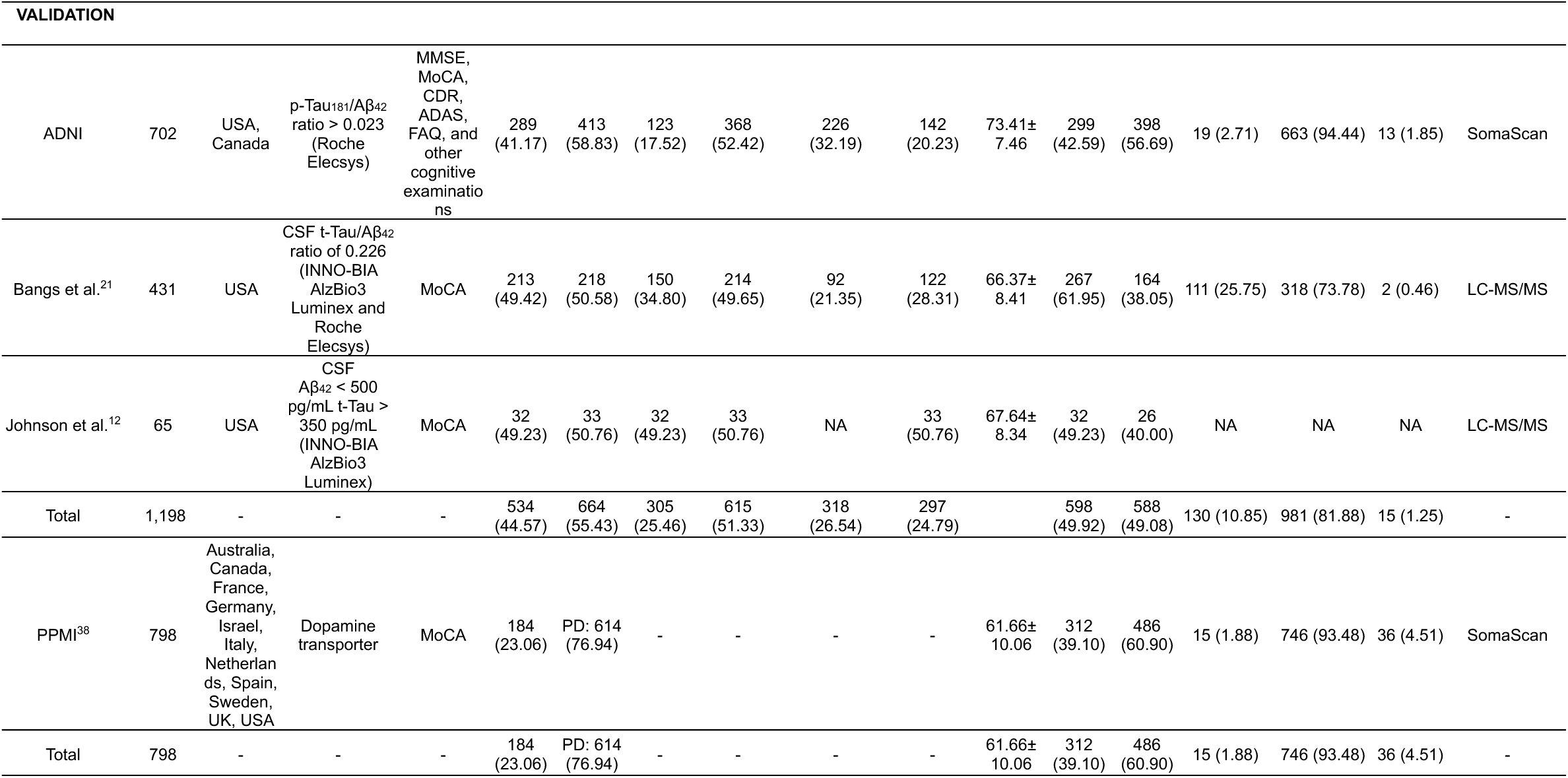
Demographic information of the selected studies.

### Biomarker discovery

Across the eight studies, 8,242 unique proteins were identified. After applying biological and statistical filters (log₂ FC ± 0.6 and *p* < 0.05) to identify differentially abundant proteins (DAPs), the list of proteins showing significant differences between AD and control groups was reduced to 1,067. Of these, only 68 proteins were shared across at least two studies (**Figure 2C**), and 41 exhibited consistent directional changes (**Figure 2D**). Proteins shared across the three test cohorts were further filtered, resulting in 13 biomarker candidates (**Figure 2E**). Down selection was performed with receiver operating characteristics (ROC) curve model, optimization indicated that an 11-protein combination provided the best performance, as inclusion of the two additional candidates did not improve diagnostic accuracy. For parsimony, this final panel was therefore defined as PPAV11 (11-protein panel for Alzheimer vote-counting). Several proteins from the PPAV11 signature have been previously reported as dysregulated in the brain and CSF of patients with AD in proteomics studies. Their molecular mechanisms and roles in disease development have also been investigated and reviewed. Frequently reported proteins include CHI3L1^4,6,14,27,30^, PKM^6,14,27,31^, NPTX2^3,4,6,14,27^, SMOC1^4,6,14,19,24,27,32^, YWHAG^6,14^_,_ YWHAZ^4,6,14,19,24,27^, and less frequently reported proteins include SPON1^6,14^, and DDAH1 ^14,33,34^. Notably, while LRRC4B^27,34^, CYCS^27,34^, and GDA^6,14^ have been reported as dysregulated in AD compared to controls, they remain less studied than the other PPAV11 proteins. Consequently, their specific mechanisms in AD remain poorly characterized. When comparing abundance levels of individual proteins between AD and control subjects (clinically and biologically diagnosed), nine (CHI3L1, LRRC4B, DDAH1, GDA, PKM, SMOC1, SPON1, YWHAG, YWHAZ) were upregulated and two downregulated (CYCS, NPTX2) in AD (**Figures S1 and S2)**. When stratifying by ethnicity, we noted a higher increase in GDA and SMOC1 protein levels in Caucasian subjects with AD (both biological and clinical diagnostic criteria) compared to African American subjects, as reported previously^23^. When stratifying by sex, we observed no clear differences (at least in two cohorts) in PPAV11 protein levels among men and women (**Figures S1 and S2**).

### Diagnosis performance

To determine diagnosis performance, two cohorts were used for model training (n = 118), three independent cohorts for validation (n = 1,198), and three additional cohorts for differential diagnosis, one of PD (n = 798) and two of A⁻T⁻ MCI (n = 488) (**Figure 3A**). Moreover, to contextualize our findings within the broader field of AD research, we compared the diagnostic and prognostic performance of PPAV11 against thirteen published proteomic signatures (**Table 2**). We found that most proteins composing all panels were unique to each signature, with only a small subset consistently identified across studies. The most frequently shared proteins among the signatures were SMOC1, YWHAG, PKM, YWHAZ, NPTX2, and GFAP. Notably, all these proteins—except GFAP—are included in PPAV11 (**Figure 3B**). This overlap reinforces the strength of our reproducibility-based approach, indicating that it aligns with proteins repeatedly associated with AD across various study designs and analytical frameworks. Conversely, the remaining proteins were either rarely observed or exclusive to individual signatures, suggesting they may be less extensively researched or more context specific.

**Figure 3.**
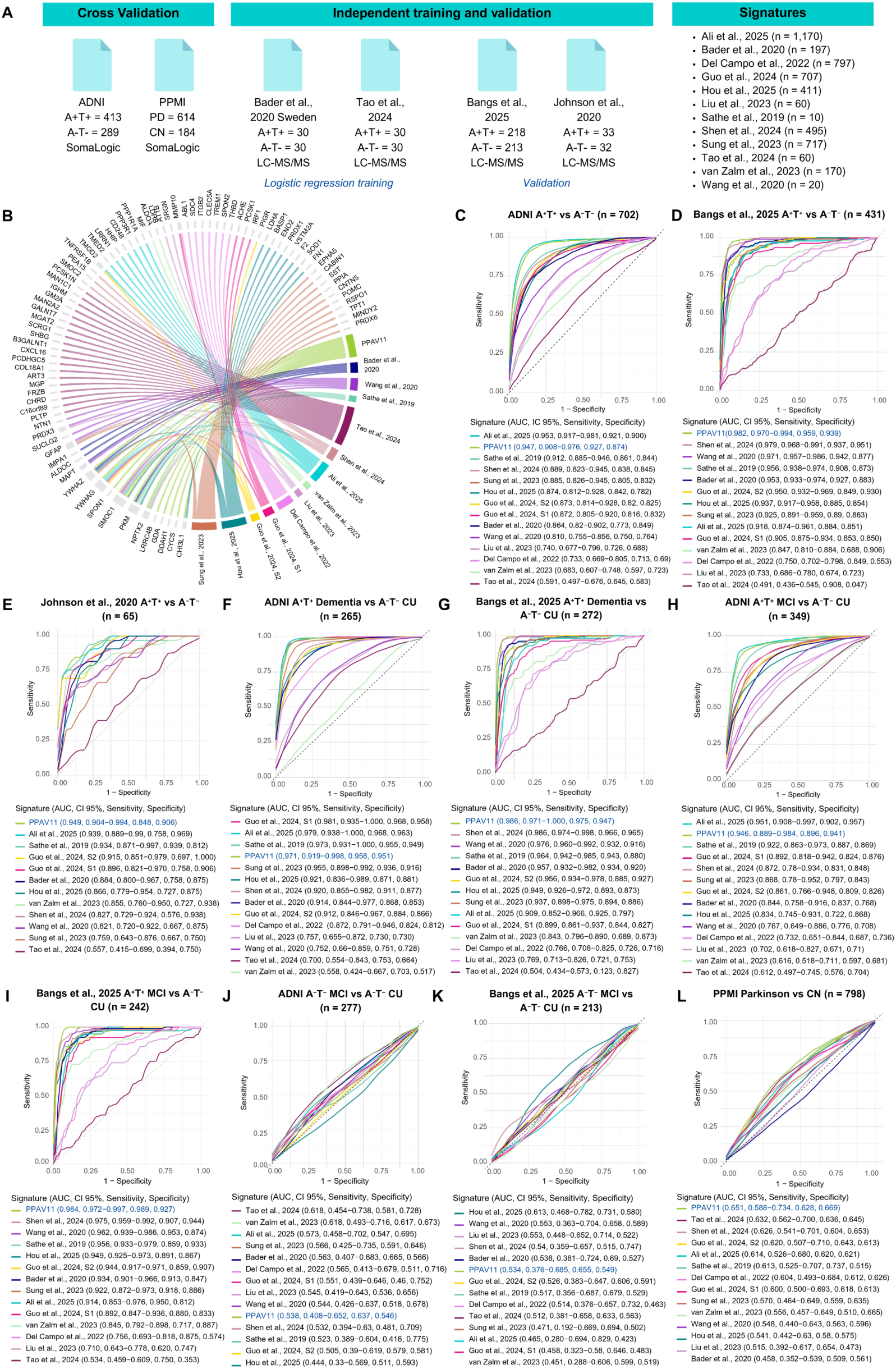
**Validation of the PPAV11 signature for Alzheimer’s disease (AD) diagnosis and benchmarking against published proteomic signatures**. (A) Overview of datasets and proteomic signatures used for validation and comparison. (B) Shared proteins between PPAV11 and the 13 comparative proteomic signatures. Receiver operating characteristics (ROC) curve analyses comparing the utility of of the signatures to distinguish between subjects stratified by (C-E) biological diagnosis (A+T+ vs A-T-); and by (F-I) biological diagnosis and clinical status (A+T+ MCI vs A-T- CU, and A+T+ dementia vs A-T- CU). Selectivity of signatures was evaluated by comparing (J-K) A-T- MCI vs A-T- CU subjects, and (L) PD vs control subjects. Sensitivity and specificity were determined by Youden’s index. Abbreviations: AD, Alzheimer’s disease; CU, cognitively unimpaired; PD, Parkinson’s Disease; CN, control; MCI, mild cognitively impaired; ROC, receiver operating characteristics; PPMI, Parkinson’s Progression Markers Initiative; ADNI, Alzheimer’s Disease Neuroimaging Initiative; LC-MS/MS, liquid chromatography coupled to Tandem mass spectrometry; CI, confidence intervals; AUC, area under the curve.

**Table 2.**
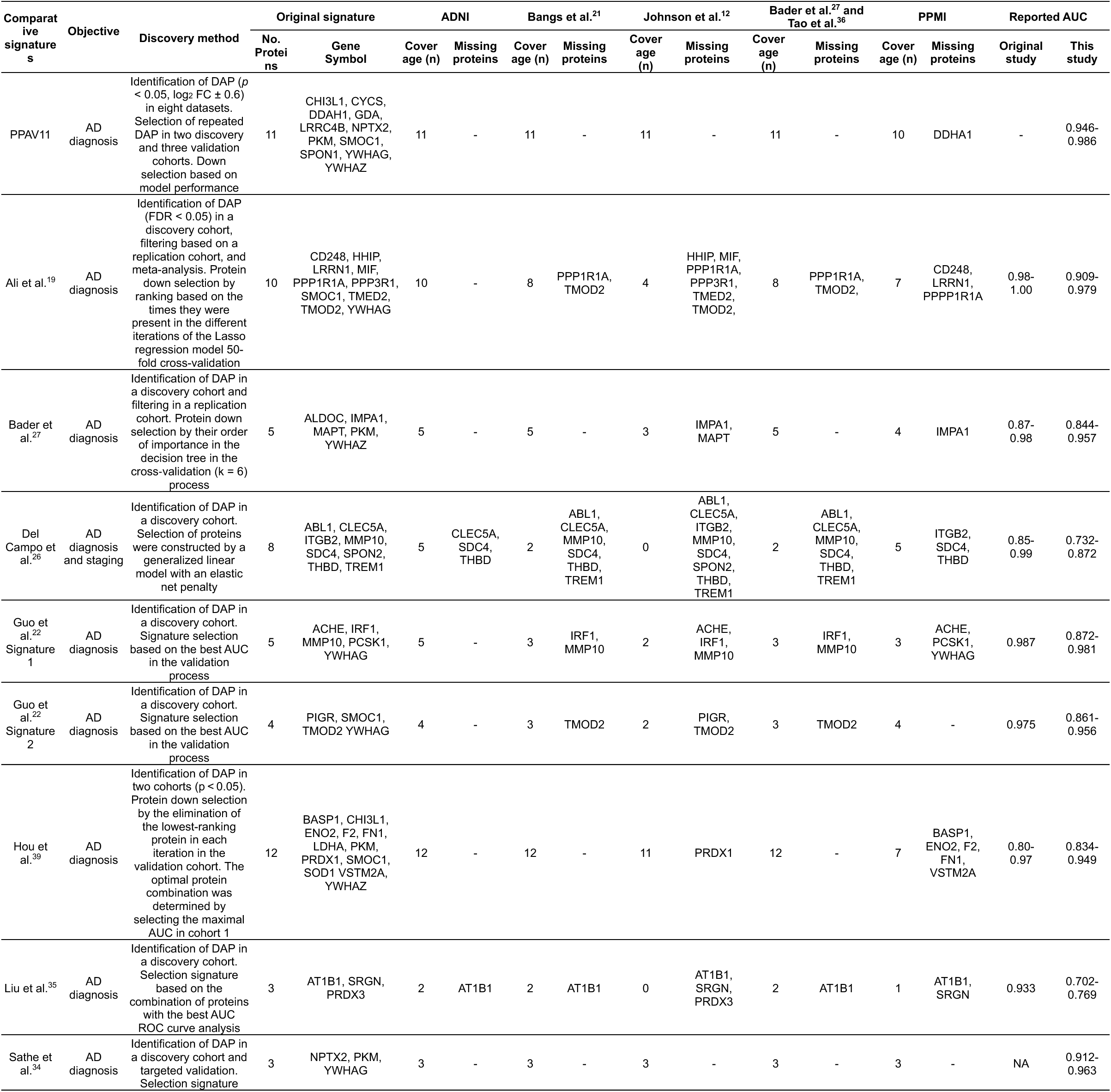
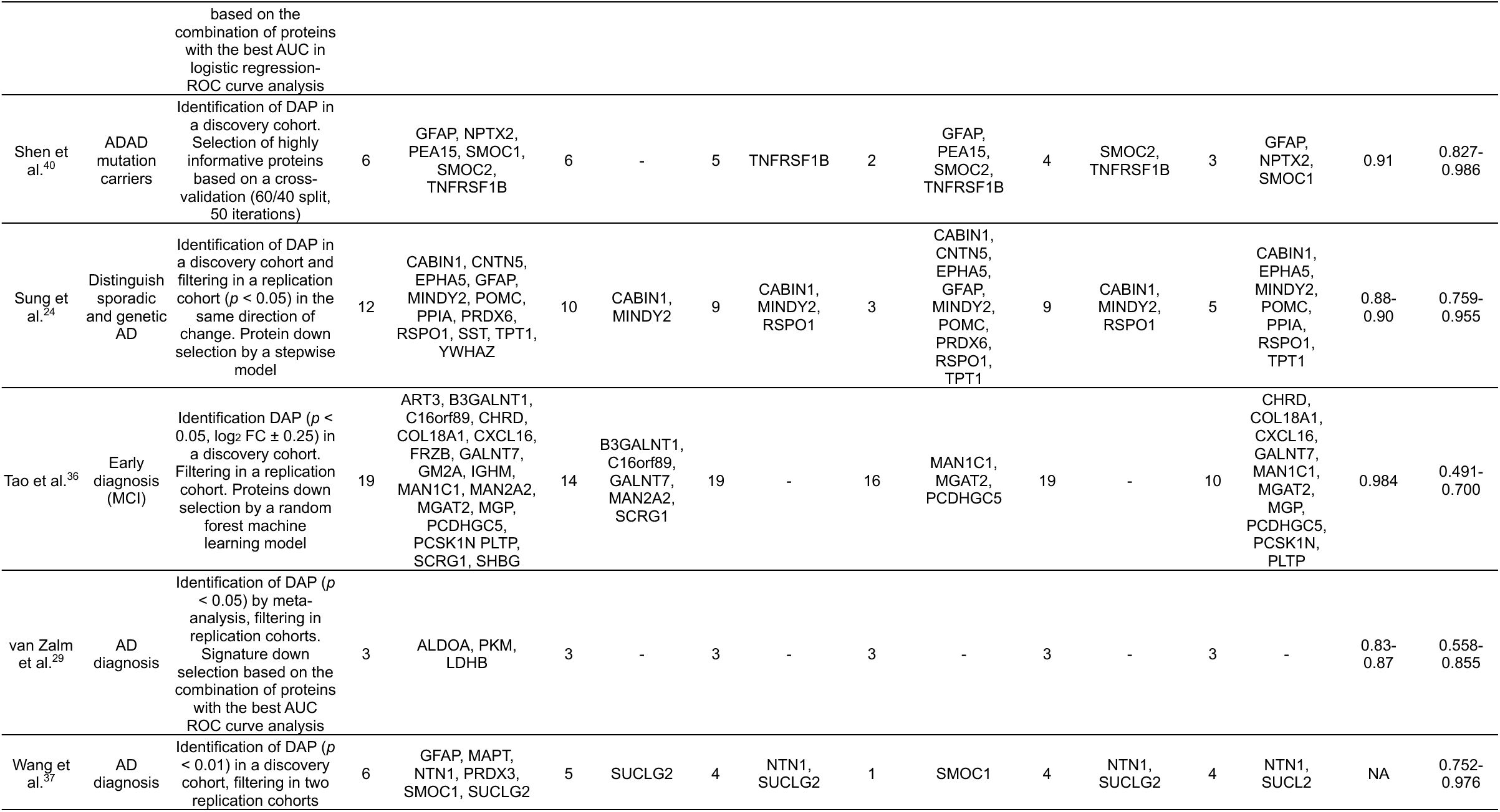
Signatures used for benchmarking.

Diagnostic performance was evaluated using two stratification frameworks: (1) a biological classification based on Aβ and p-Tau biomarkers (A⁺T⁺ vs A⁻T⁻, and A⁺T⁻ vs A⁻T⁻)^1^, and (2) a combined biological (A⁺T⁺ vs A⁻T⁻) and clinical diagnostic classification (CI vs cognitively unimpaired [CU])^2^. We first performed principal component analysis (PCA) as an exploratory analysis to evaluate whether PPAV11 could distinguish subjects with and without AD. Across the three validation cohorts, significant clustering differences were observed under both biological and clinical stratifications (MANOVA Pillai’s trace, *p*<0.05; **Figures S3A–I**). In contrast, minimal or no separation was observed when comparing A⁻T⁻ MCI subjects from controls (**Figures S3J and S3K**) or when comparing individuals with and without PD (**Figure S3L**). These findings suggested both effective separation between AD and control groups and a selectivity for AD. To quantitatively evaluate diagnostic performance across cohorts, we next performed ROC curve analysis. Across independent validation cohorts, PPAV11 demonstrated stable diagnostic performance, yielding consistent area under the curve (AUC) values across both biological and clinical classifications (**Figures 3C-I)**. Notably, PPAV11 achieved the highest AUC values among the evaluated signatures in two independent validation cohorts, Bangs et al.^21^ and Johnson et al.^12^, under biological stratification (AUC = 0.982 and 0.949, respectively; **Figures 3D and 3E**). High diagnostic accuracy was also observed under the combined biological and clinical framework in the Bangs cohort, with AUC values of 0.986 for dementia (**Figure 3G**) and 0.984 for MCI (**Figures 3I**). Consistent performance was observed in the ADNI cohort for both biological stratification (AUC = 0.947; **Figure 3C**) and the combined biological and clinical classification, for both dementia (AUC = 0.971; **Figure 3F**) and MCI (AUC = 0.946; **Figure 3H**), supporting the robustness of PPAV11 acorss datasets. Although some signatures achieved slightly higher AUC values within ADNI, their performance ws less consistent across external cohorts, suggesting lower generalizability. When stratified solely by amyloid status (A⁺ vs A⁻), performance was highest using amyloid-PET classification (AUC = 0.906) and remained comparable to previously published panels under CSF-based stratification (**Figures S4A and S4B**). When evaluating differential diagnostic performance in A⁻T⁻ MCI and PD groups, none of the evaluated signatures achieved high diagnostic accuracy in these non-AD groups (AUC < 0.622), suggesting selectivity for AD (**Figure 3J-L**). Analysis of the individual proteins comprising PPAV11 revealed variability in diagnostic performance across cohorts and stratification schemes (**Figure S5**), highlighting the increased stability conferred by the multi-protein signature across cohorts and disease stages. Importantly, comparative analysis showed that the AUC values obtained in our study were largely consistent with those reported in the original publications (**Table 2**), except for Liu et al.^35^ and Tao et al.^36^. This concordance across independent studies supports the reproducibility of our modeling framework and the robustness of the comparative evaluation. Observed discrepancies are most likely attributable to differences in protein coverage across cohorts, diagnostic stratification, and modeling strategies across studies. While several previously reported panels exhibited broader variability across independent datasets, PPAV11 demonstrated stable performance across cohorts with balanced sensitivity and specificity, supporting the robustness of the data integrative and reproducibility-based strategy used to derive this signature.

### Prognosis performance

For prognostic validation, we analyzed longitudinal data from the ADNI cohort, as it provided sufficient follow-up information to evaluate cognitive progression. We focused on two clinically relevant transitions: CU individuals who progressed to A⁺T⁺ MCI (n = 141), and A⁺T⁺ MCI individuals who progressed to A⁺T⁺ dementia (n = 174). Participants were required to have at least two clinical visits, with follow-up durations ranging from 6 months to 12.5 years. To assess prognostic performance, individuals were stratified into high- and low-score groups for each signature, and the relative risk of cognitive decline was estimated between groups. In addition to published proteomic signatures, we included established AD core biomarker ratios and individual protein levels that have been studied with AD-related cognitive decline^41–44^. For the transition from CU to A⁺T⁺ MCI, core AD biomarker ratios—including t-Tau/Aβ42 (Hazard Ratio [HR] = 5.85, *p* = 0.002) and p-Tau/Aβ42 (HR = 6.11, *p* = 0.001)—as well as Aβ42 alone (HR = 10.55, *p* = 0.002), showed the strongest predictive performance (**Figure 4A**). This result was anticipated given that the classification partially depends on alterations in these biomarkers. The proteomic panel reported by Sung et al.^24^, also showed strong predictive capacity within this dataset (HR = 27.72, *p* = 0.001). Among the evaluated signatures, PPAV11 demonstrated outstanding prognostic performance, with individuals in the high-score group exhibiting an approximately five-fold increased risk of progression (HR = 4.96, *p* = 0.004) compared to those in the low-score group (**Figure 4B**). The transition from A⁺T⁺ MCI to A⁺T⁺ dementia revealed a distinct pattern. In this later stage of disease progression, core AD biomarkers were not the strongest predictors of clinical conversion (**Figure 4C**). Instead, a panel reported by Guo et al.^22^ showed the highest predictive performance (HR = 3.56, *p* = 2.42 ×10^-7^). PPAV11 also demonstrated substantial prognostic capacity in this transition, with individuals in the high-score group exhibiting an approximately three-fold increased risk of progression (HR = 3.23, *p* = 3.13×10^-17^) to dementia compared to those with low scores (**Figure 4D**). Taken together, these results indicate that different biomarker classes capture complementary aspects of AD progression. While canonical AD biomarkers remain strong predictors of early pathological transitions, multi-protein signatures can capture additional biological variation associated with later disease stages. Importantly, since some of the previously reported signatures were derived from the ADNI cohort, PPAV11 provided the strongest prognostic signal under fully independent evaluation in both transitions, highlighting the robustness and generalizability of the reproducibility-based approach.

**Figure 4.**
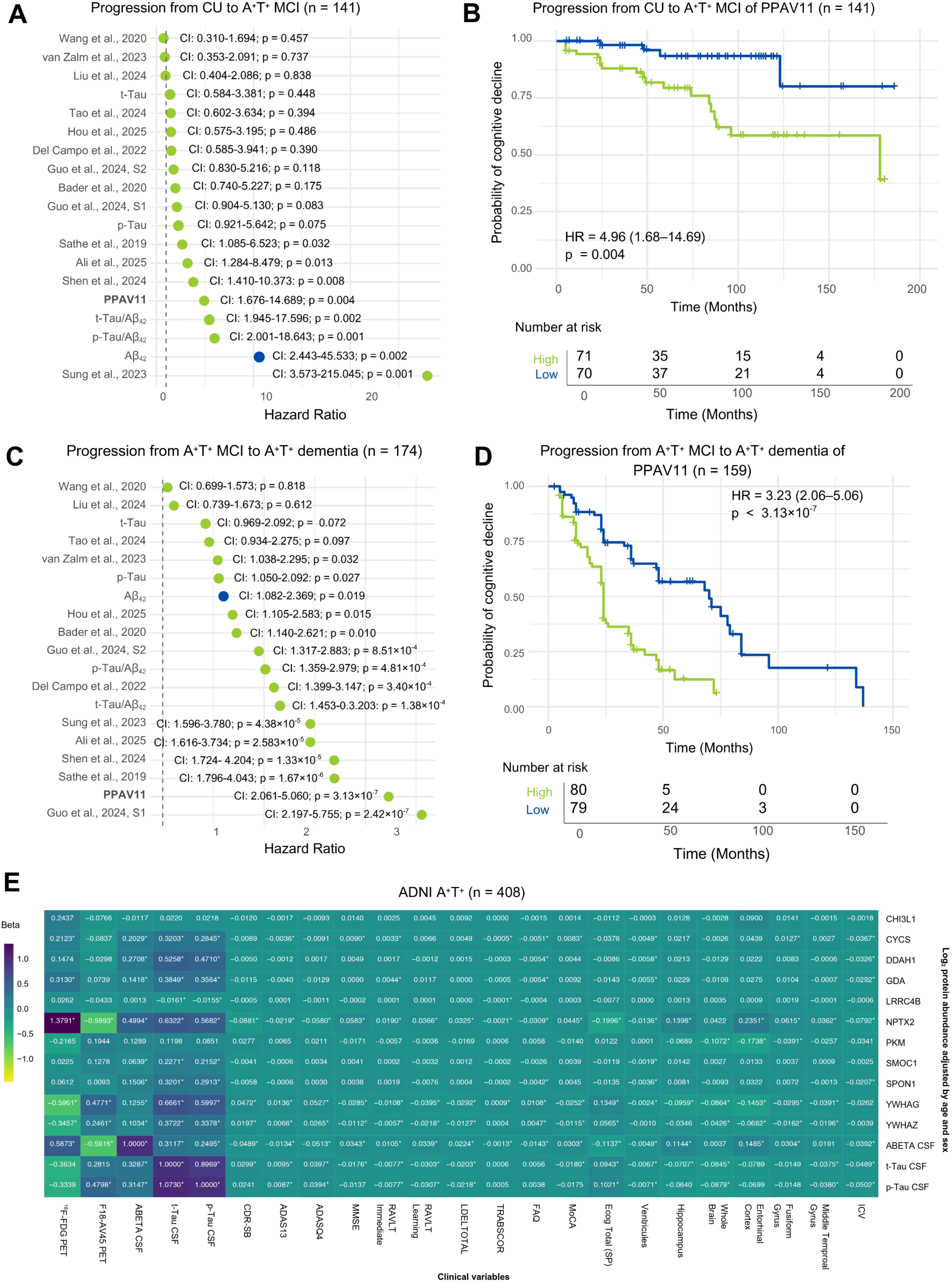
Prognostic validation of PPAV11 for cognitive decline and correlation analysis of PPAV11 proteins with clinical variables in the ADNI cohort. Cox proportional hazards model comparing progression from (A) CU to A^+^T^+^ MCI and from (C) A^+^T^+^ MCI to A^+^T^+^ dementia using binary scores (low and high) adjusted for age and sex. (B, D) Kaplan–Meier survival curves comparing cognitive decline between the PPAV11 low and high groups. *P-values* were calculated using Cox proportional hazards regression. (E) Heatmap shows correlations of individual PPAV11 proteins with cognitive tests, brain volume measurements, and biological biomarkers. Beta coefficient values were derived from LIMMA analysis and represent the association of change in protein abundance (log_2_) per unit of each clinical variable. Models were adjusted for age and sex. Volume metrics are expressed in cm^2^. Significance (adjusted-p < 0.05, Benjamini-Hochberg correction) is highlighted by an asterisk (*). Abbreviations: AD, Alzheimer’s disease; CU, cognitively unimpaired; MCI, mild cognitively impaired; ADNI, Alzheimer’s Disease Neuroimaging Initiative; ^18^F-FDG PET, ^18^F-fluorodeoxyglucose positron emission tomography; F18-AV45 PET, Florbetapir F18 positron emission tomography; t-Tau, total Tau; p-Tau, phosphorylated Tau; CDR-SB, Clinical Dementia Rating–Sum of Boxes; ADAS, Alzheimer’s Disease Assessment Scale; MMSE, Mini Mental State Examination; RAVLT, Rey Auditory Verbal Learning Test; LDEDLTOTAL, logical memory delayed recall total; TRABSCOR, Trail Making Test; FAQ, Functional Activities Questionnaire; MoCA, Montreal Cognitive Assessment; ECOG (SP) Everyday Cognition, spouse report; ICV, intracerebroventricular.

### Correlation analysis of PPAV11 with clinical variables

To evaluate the clinical relevance of PPAV11, we performed a LIMMA correlation analysis using data from the ADNI cohort (n = 408, A⁺T⁺ subjects with complete clinical information), which provided the most comprehensive set of clinical and imaging variables. We computed correlations for each protein in the signature, as well as for CSF Aβ42, total Tau (t-Tau), and p-Tau (derived from Elecsys immunoassay), against measures of cognitive performance, regional brain volumes, CSF- and PET-based biomarkers, adjusting by age and sex (**Figure 4E**). Among the PPAV11 proteins, NPTX2 exhibited the strongest and most consistent associations with clinical measures of cognitive decline and neurodegeneration, in line with previous reports linking NPTX2 to cognitive impairment in AD^45–47^. NPTX2 showed significant associations across all cognitive tests, including the Clinical Dementia Rating Sum of Boxes (CDR-SB; β = –0.088, *adjusted-p* = 2.4×10^-13^), the Alzheimer’s Disease Assessment Scale–Cognitive Subscale (ADAS13; β = –0.021, *adjusted-p* = 1.1×10^-13^), the Mini–Mental State Examination (MMSE; β = 0.058, *adjusted-p* = 3.7×10^-13^), the Functional Activities Questionnaire (FAQ; β = –0.039, *adjusted-p*= 6.8×10^-17^), and the Montreal Cognitive Assessment (MoCA; β = 0.0445, *adjusted-p* = 3.05×10^-8^). In addition, NPTX2 was strongly associated with structural measures of neurodegeneration, including hippocampal (β = –0.139, *adjusted-p* = 6.02×10^-5^) and entorhinal cortex (β = –0.235, *adjusted-p* = 3.90×10^-7^) volume as well as ventricular expansion (β = –0.013, *adjusted-p* = 2.6×10^-24^) and metabolic activity measured by ¹⁸F-FDG-PET (β = 1.37, *adjusted-p* = 1.1×10^-12^) (**Figure 4E**). Consistent with prior studies^3,46,48,49^, lower NPTX2 levels were associated with worse cognition and greater atrophy in hippocampal and entorhinal regions—key structures affected during AD progression^50–52^. Beyond NPTX2, several additional proteins in the PPAV11 signature showed notable associations with neurodegenerative imaging markers. YWHAG and YWHAZ exhibited strong correlations with regional brain atrophy, particularly across temporal and limbic structures including the fusiform gyrus, entorhinal cortex, middle temporal gyrus (MTG), hippocampus, and whole brain volume. When effect sizes were scaled for direct comparison (**Figure S6**), correlations between NPTX2, YWHAG and YWHAZ and structural atrophy measures exceeded those observed for core CSF biomarkers in several regions. Similarly, these proteins showed strong associations with metabolism measured by ¹⁸F-FDG-PET, exceeding those observed for t-Tau and p-Tau. For amyloid pathology measured by AV45 PET imaging, NPTX2 and YWHAG demonstrated associations comparable to those observed for CSF Aβ_42_. Notably, several PPAV11 proteins showed significant associations with Tau pathology: CYCS, DDAH1, SMOC1, SPON1, YWHAG, and YWHAZ all correlated strongly with t-Tau and p-Tau levels, capturing Tau-linked molecular processes. These findings highlight the signature’s clinical relevance—especially NPTX2 and YWHAG in tracking cognitive decline—outperforming core CSF biomarkers across cognitive and imaging measures. While Aβ and Tau define AD pathology, NPTX2 and YWHAG better reflect downstream synaptic changes tied to symptom expression. These results further support the value of multi-protein signatures such as PPAV11 for integrating complementary molecular signals across the AD pathological cascade.

### Brain cell-type enrichment analysis and regional dysregulation

To determine the brain regions in which PPAV11 proteins are dysregulated in AD compared with control subjects, we queried the NeuroPro platform^53^. This resource compiles dysregulated proteins (FDR < 0.05 or *p* < 0.05 with FC ≥ 1.5 or ≤ 0.667) across 32 proteomic studies covering 13 brain regions. Among PPAV11 proteins, the highest proportion of dysregulated proteins was observed in the frontal cortex, hippocampus, and parietal cortex (**Figure 5A**), regions whose dysregulation aligns with the characteristic atrophy pattern of typical AD^54–56^. To further explore the cellular landscape of PPAV11 proteins, we used single-nucleus transcriptomics data from the Comparative Viewer^57^ in the Brain Knowledge Platform (https://brain-map.org/bkp) from the Allen Institute. First, we examined their gene expression patterns using data from the MTG of five neurotypical donors^57^. SMOC1 and SPON1 were strongly enriched in non-neuronal populations—specifically oligodendrocyte progenitor cells (OPCs) and astrocytes, respectively (**Figure 5B**). These proteins were also detectable in GABAergic neurons, although at lower levels, and showed minimal expression in glutamatergic neurons. In contrast, DDAH1, GDA, PKM, YWHAG, YWHAZ, and LRRC4B showed relatively uniform enrichment in glutamatergic neurons compared with GABAergic neurons, while several of these proteins were broadly expressed across GABAergic neuronal subtypes. CHI3L1 showed low expression in glutamatergic neurons and astrocytes. Overall, PPAV11 proteins were expressed across both neuronal and non-neuronal populations, and some displayed strong cell-type specificities. We next examined PPAV11 gene expression dynamics across AD pseudoprogression in the SEA-AD cohort, which integrates quantitative neuropathology from 84 donors stratified by pseudoprogression score (CPS; higher values indicate greater disease severity). We identified cell-type-specific correlations and distinct trajectories between PPAV11 expression and AD severity (**Figure 5C**). In GABAergic neurons (Vip, Sst, Pvalb, and Sncg), CYCS, GDA, LRRC4B, PKM, and YWHAZ showed progressive expression decreases with increasing CPS, whereas YWHAG followed a U-shaped trajectory. In glutamatergic L2/3 IT neurons, GDA, NPTX2, and YWHAZ decreased with disease severity, while LRRC4B and PKM increased. In glial populations, protoplasmic astrocytes showed increased CHI3L1 and decreased SPON1 expression across pseudoprogression. In myelinating oligodendrocytes and OPCs, NPTX2 expression increased, whereas PKM expression increased in disease-associated microglia (DAM). Notably, several PPAV11 genes exhibited expression changes that temporally coincided with the emergence of amyloid plaques and Tau tangles. We therefore focused on these transcriptional shifts as they occur at key stages of AD pathological progression. NPTX2 alterations in L2/3 IT neurons may reflect disruptions in synaptic plasticity and neuronal assemblies^46,58^, whereas its expression changes in OPCs and oligodendrocytes have not been previously described in AD. PKM overexpression in DAM has been linked to metabolic reprogramming^59^, while changes in CHI3L1 expression in astrocytes are associated with shifts toward reactive and inflammatory glial states^30,60–62^. Together, the temporal alignment of these transcriptional changes with amyloid and tau pathology suggests that PPAV11 genes capture key biological transitions associated with AD progression.

**Figure 5.**
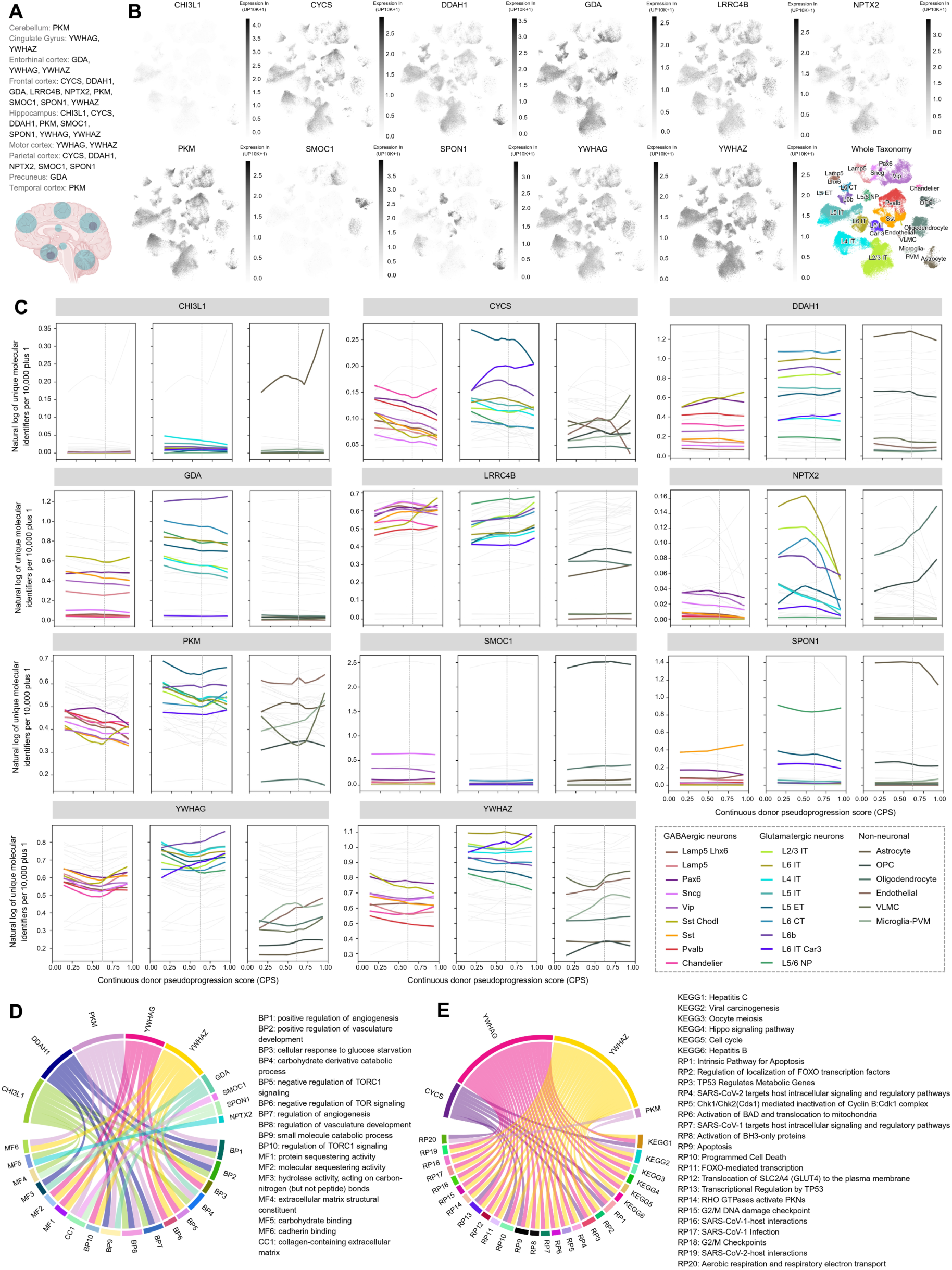
Bioinformatic analysis of PPAV11 using enrichment and single-nucleus RNA sequencing (snRNA-seq) expression databases. (A) Brain regions with dysregulated PPAV11 proteins in AD vs control subjects in NeuroPro proteomics database (https://neuropro.biomedical.hosting/). (B) Comparative expression of PPAV11 protein members across brain cell types (snRNA-seq) in five neurotypical donors from the MTG reference cohort and (C) expression changes of PPAV11 proteins across disease pseudoprogression in 84 donors from the SEA-AD cohort, in the transcriptomics comparative viewer (https://knowledge.brain-map.org/data/5IU4U8BP711TR6KZ843/2CD0HDC5PS6A58T0P6E/compare) from Allen Institute. Enrichment analyses (D) Gene Ontology (GO) and (E) REACTOME databases are shown for terms with FDR < 0.05. Abbreviations: AD, Alzheimer’s disease; snRNA-seq, single-nucleus RNA sequencing; SEA-AD, Seattle Alzheimer’s Disease Brain Cell Atlas; MTG, Middle temporal gyrus. Brain image from panel A was created with biorender.com.

### Functional enrichment analysis

To explore the biological processes captured by the PPAV11 proteins, we performed enrichment analyses using the GO^63^ and REACTOME^64^ databases. Enriched GO terms were primarily associated with angiogenesis (FDR = 0.025–0.040), carbohydrate metabolism (FDR = 0.025–0.040), and TOR signaling regulation (FDR = 0.025–0.047) (**Figure 5D**). Pathway analysis using REACTOME further identified enrichment for immune and inflammatory responses (FDR = 0.0007–0.036), apoptosis (FDR = 0.0007–0.034), metabolic pathways (FDR = 0.0007–0.045), and transcription factor regulation (FDR = 0.0007–0.013) (**Figure 5E**), highlighting the involvement of PPAV11 proteins in multiple biological processes widely implicated in AD pathology^8,19,65–68^. Among these pathways, alterations in glucose metabolism—represented by proteins such as GDA, PKM, and YWHAZ—have been consistently reported as one of the most prominent disruptions in AD proteomic studies^12,14,27,29^. Dysregulation of glycolytic enzymes has been linked to impaired ATP production and reduced neuronal firing, ultimately compromising synaptic maintenance and neuronal function^29,69^. Consistent with this, enrichment of electron transport pathways and apoptosis signaling, further supports the presence of mitochondrial dysfunction, a hallmark of AD associated with impaired oxidative phosphorylation and increased reactive oxygen species production^37^. Immune activation and inflammation also emerged as a central axis within PPAV11. Microglia have been associated with both protective and neurodegenerative roles ^70^. While early activation is linked to Aβ clearance, chronic activation is thought to sustain a proinflammatory environment that contributes to synaptic dysfunction and neuronal loss^71–74^. Consistent with this, large-scale proteomic studies have repeatedly identified glial-associated modules—including proteins of PPAV11 such as SMOC1, SPON1, and CHI3L1—linked to immune signaling, oxidative stress, morphogenesis, and angiogenic processes^4,12^. Angiogenesis and vascular-related processes were also enriched within PPAV11, involving proteins such as CHI3L1, DDAH1, and PKM. Increasing evidence suggests that vascular dysregulation and impaired angiogenic signaling contribute to AD pathology by promoting hypoxia, blood–brain barrier dysfunction, and reduced endothelial clearance of Aβ^75,76^. In addition, proteins involved in mTOR signaling—including members of the 14-3-3 family such as YWHAG and YWHAZ—were represented within the enriched pathways. Dysregulation of mTOR has been linked to multiple processes, including impaired autophagy, facilitation of the accumulation of misfolded proteins, mitochondrial dysfunction, neuroinflammation, and vascular dysfunction^69,74,77^. Collectively, these results indicate that PPAV11 captures key mechanisms underlying AD pathophysiology, spanning metabolic stress, neuroinflammation, vascular remodeling, mitochondrial dysfunction, and synaptic regulation.

## Discussion

Over the past decade, CSF proteomics studies have identified numerous candidate biomarkers for AD diagnosis, expanding our understanding of the molecular processes underlying neurodegeneration. These efforts have highlighted the biological heterogeneity of AD and the involvement of diverse pathways beyond the core amyloid and Tau pathologies, emphasizing the need for biomarker signatures that better capture disease complexity and its clinical manifestations. At the same time, the growing number of proteomic datasets has generated a large and partially overlapping landscape of candidate biomarkers, raising challenges for integration and reproducibility across cohorts and analytical platforms. In particular, many proposed signatures have been derived using distinct discovery strategies and evaluated in different cohorts, limiting direct comparisons of their relative performance and robustness. To address this gap, in this study, we implemented a reproducibility-driven prioritization framework to identify CSF proteins that consistently discriminate AD across independent cohorts and proteomic platforms. Rather than optimizing peak performance within a single dataset, we applied biologically informed effect-size thresholds, statistical support, and directional concordance across cohorts to minimize context-specific signals and prioritize stable molecular alterations. This strategy led to the identification of an 11-protein panel (PPAV11) that demonstrated robust performance after rigorous validation and systematic benchmarking against published protein signatures. Because PPAV11 was derived from reproducible signals across independent studies, many of its constituent proteins have been previously reported in AD and extensively characterized at the molecular level. Among them, SMOC1 is the most consistently described^4,14,19,24,27,32,65^, having been repeatedly associated with core AD pathology and shown to colocalize with both Aβ plaques and p-Tau tangles^32^, supporting its role as an early biomarker. CHI3L1 has emerged as a selective astrocytic marker indicative of reactive and neurotoxic inflammatory states^62,78,79^. In parallel, YWHAG and YWHAZ have been linked to Tau-related pathology, while together with NPTX2, they reflect alterations in synaptic maintenance and circuit integrity^3,46,58^. On the other hand, some proteins of PPAV11 such as LRRC4B^27,34^, CYCS^35,37^, and GDA^6,14^ have been reported as dysregulated in AD compared to control subjects, though they are still relatively underexplored. To evaluate the diagnostic performance of PPAV11, we compared it with previously published CSF proteomic signatures^19,22,24,26,27,29,34–37,39,40^ using the same validation cohorts and analytical framework. This head-to-head benchmarking enabled direct assessment of the relative performance of each signature under the same conditions. Across independent validation datasets, PPAV11 demonstrated the most stable and consistently strong diagnostic performance (AUC range 0.946–0.986). Its robustness was observed across biological and clinical diagnostic stratifications, early and late disease stages, CSF- and PET-based classifications, and distinct proteomic platforms. In addition, PPAV11—and all signatures—showed disease selectivity, as diagnostic performance was markedly reduced when evaluated in A⁻T⁻ MCI and in PD cohorts. While some previously reported panels achieved high AUC values in specific datasets, their performance varied considerably when evaluated across cohorts and diagnostic stratifications. Differences in the proteomic platforms used across validation cohorts likely contributed to some of the observed variability. Signatures derived from a given platform may exhibit decreased performance when evaluated using data generated by a different technology, due in part to reduced protein coverage. In contrast, PPAV11—derived from LC-MS/MS and Olink datasets—maintained consistent protein coverage and strong discrimination in the SomaScan-derived cohort, with minimal inter-cohort and inter-platform variability. Together, these findings highlight that peak performance within a single dataset does not necessarily translate into cross-context generalizability, emphasizing cross-cohort and cross-platform stability as important criteria for the development of clinically transferable biomarker signatures. Importantly, the capacity of our framework to reproduce the performance of most previously published signatures further supports the robustness of the comparative analyses. Remaining discrepancies across studies likely reflect differences in the selected cohorts, protein coverage, diagnostic stratification and modeling strategies. Prognostic analyses further highlighted the relevance of PPAV11 across disease stages. Among the evaluated protein signatures, only a limited subset showed statistically significant associations with disease progression, and most were informative primarily at later stages. In contrast, PPAV11 demonstrated significant associations across both key clinical transitions—from CU to A⁺T⁺ MCI and from A⁺T⁺ MCI to A⁺T⁺ dementia—outperforming the majority of previously published panels. As the observed HR exceeded previously reported estimates^19,42^, these findings should be interpreted cautiously given potential cohort-specific effects. During the CU to A⁺T⁺ MCI transition, core AD biomarkers showed the strongest predictive performance. Importantly, as Aβ and Tau positivity are required to define this transition, it can influence their prognostic capacity in this context. Nevertheless, PPAV11 demonstrated performance comparable to established CSF ratios used for AD classification^1,80^, suggesting that the molecular processes captured by this proteomic signature are relevant during early stages of disease progression. In contrast, during the transition from A⁺T⁺ MCI to A⁺T⁺ dementia, PPAV11 showed the strongest prognostic performance among signatures derived from independent discovery cohorts, exceeding that of core AD biomarkers. This observation is consistent with accumulating evidence that amyloid and Tau pathology alone do not fully explain the trajectory of cognitive decline in AD^81,82^. In this context, the capacity of PPAV11 to capture risk across both transitions suggests that the biological pathways represented by the signature may reflect broader mechanisms contributing to clinical progression beyond Aβ and Tau. The prognostic capacity of PPAV11 may partly reflect the biological relevance of its constituent proteins, several of which show significant associations with clinical and neuroimaging measures. Among these, NPTX2 displayed the strongest and most consistent correlations across outcomes, with lower NPTX2 levels associated with worse cognitive performance and greater brain atrophy. When directly compared with core AD biomarkers, NPTX2 showed stronger associations with multiple cognitive and functional measures, including CDR-SB, MoCA, MMSE, and ADAS scores. Regarding neurodegeneration, NPTX2, together with YWHAG, and YWHAZ also showed stronger correlations with regional brain atrophy than core CSF biomarkers in several structures closely linked to disease progression, including the hippocampus and entorhinal cortex^50,83,84^. These findings are consistent with previous reports highlighting the relevance of these proteins in the clinical manifestations of AD^3,46,85,86^ and align with growing evidence that synaptic proteins are among the biomarkers most strongly associated with cognitive decline^87–90^. Previously, NPTX2 has been proposed as a sensitive marker of early circuit dysfunction and cognitive decline^3,46^, likely reflecting its role in maintaining excitatory–inhibitory balance, synaptic homeostasis, and neuronal plasticity^58^ Meanwhile, 14-3-3 family proteins (including YWHAG and YWHAZ) have been proposed as key regulators of synaptic transmission and plasticity^91^. Their regulatory roles have been primarily characterized in neurodevelopment^92,93^; however, their consistent alteration across multiple conditions—including neurodegenerative diseases and epilepsy—suggests a broader involvement in synaptic dysregulation. The biological diversity of PPAV11 is further supported by convergent evidence across multiple molecular layers linking CSF alterations to brain pathology. Regional brain proteomic datasets consistently showed dysregulation of PPAV11 proteins in the frontal, hippocampal, and parietal cortices, in agreement with our LIMMA correlation results linking PPAV11 to structural brain alterations. Complementary single-nucleus transcriptomic brain analyses further indicated that these genes are expressed across both neuronal and glial populations, consistent with the multicellular nature of AD^3,32,78^. AD pseudoprogression analyses revealed that several PPAV11 genes exhibit expression trajectories that correlate with disease severity at the cellular level. Some of the trajectories temporally align with the emergence of amyloid plaques and Tau tangles, suggesting that the molecular processes captured by PPAV11 may reflect key biological transitions associated with disease progression rather than static pathological states. Those include expression changes in NPTX2 within glutamatergic neurons, CHI3L1, PKM and SPON1 in protoplasmic astrocytes and PKM in DAM. Together, these findings suggest that PPAV11 reflects disruptions in cell-type composition and molecular processes in the AD brain, including impaired synaptic plasticity and neuronal circuit imbalance^94,95^ as well as activation of AD-associated glial subpopulations linked to inflammatory responses and neurodegeneration^74,96,97^. Collectively, the observed cellular and transcriptional trajectories point to coordinated alterations across multiple biological systems, capturing progressive disease-related transitions. Consistent with these findings, functional enrichment analyses revealed coordinated involvement of PPAV11 proteins across multiple biological pathways—including angiogenesis, glucose metabolism, mTOR signaling, immune activation, inflammation, apoptosis, and transcriptional regulation—repeatedly implicated in AD by multi-omics studies^4,10,12,19,27^. Complementing these observations, longitudinal CSF proteomic studies in autosomal dominant AD^6^, Down syndrome–associated AD^98^, and sporadic AD^14^ consistently report stage-dependent changes in several PPAV11 proteins. SMOC1 and SPON1 proteins appear among early changes occurring decades before symptom onset, followed by alterations in metabolic regulators such as PKM and GDA, immune-associated proteins including CHI3L1, and later synaptic markers such as NPTX2. The convergence of these temporally ordered trajectories across genetically distinct and sporadic forms of AD, together with the multi-layered evidence presented here, supports the notion that PPAV11 captures biologically staged processes that unfold across the disease continuum. Importantly, the temporal alignment of these molecular changes with the emergence of Aβ plaques and Tau tangles—while also extending to intermediate and symptomatic stages—suggests that this signature reflects dynamic transitions associated with the progression from early pathology to downstream clinical manifestations. Thus, these findings reinforce PPAV11’s relevance for stage-aware stratification and highlight its potential utility in longitudinal and interventional settings.

Despite these strengths, several limitations should be acknowledged. Although we assembled a large cohort for discovery and diagnostic validation, analyses related to disease progression and correlation with clinical variables were restricted to a single cohort, limiting the assessment of cross-cohort and cross-platform reproducibility. Thus, future studies leveraging longitudinal data across independent cohorts will be essential to establish the generalizability of these findings. In addition, although our benchmarking framework proved stable and reproducible, not all previously published signatures could be fully assessed due to incomplete protein coverage across datasets, particularly for panels derived from SomaScan or Olink in the LC-MS/MS validation cohorts. Finally, the predominance of cohorts derived from North American and European populations may limit broader applicability, underscoring the need for validation across more diverse populations. In summary, this study highlights the value of reproducibility-centered strategies for biomarker discovery in heterogeneous neurodegenerative diseases, as well as the importance of rigorous cross-cohort and cross-platform validation alongside deep biological integration. Through this approach, our framework prioritizes signatures that are not only performant but also robust and biologically grounded. Our findings further support the advantage of protein panels or signatures in capturing complementary dimensions of disease biology—including cellular heterogeneity, temporal dynamics, and clinical variability— thereby enhancing diagnostic stability, prognostic utility, and patient stratification. More broadly, such integrative approaches may help reduce fragmentation in the field, accelerate the identification of clinically translatable biomarkers for disease staging and monitoring, and support the design of biologically grounded clinical trials. Finally, this work underscores the critical role of open-access omics resources, illustrating how data sharing enables scalable and reproducible strategies to refine and prioritize molecular candidates for translational applications.

## Data Availability

Proteomics data from the ADNI cohort are available at https://adni.loni.usc.edu. Proteomics datasets from Bangs et al.^21^ and Johnson et al.^12^ are available at https://www.synapse.org/, under project IDs syn65461849 and syn20821165, respectively. Proteomics data from the PPMI^38^ cohort are available at www.ppmi-info.org. Additional datasets used in this study are provided in the supplemental information of the corresponding articles. The original code supporting this work is available in https://zenodo.org/records/19687369.

## Resource availability

### Lead contact

For further information or resource requests, please contact the lead investigator, Dr. Aldo Moreno-Ulloa.

### Materials availability

This study did not generate new materials.

## Acknowledgments

We thank the participants and their families for generously contributing their data, all contributors to the acquisition and processing of the datasets, and the researchers who made these datasets available in the context of open science. Data collection and sharing for the Alzheimer’s Disease Neuroimaging Initiative (ADNI) is funded by the National Institute on Aging (National Institutes of Health Grant U19 AG024904). The grantee organization is the Northern California Institute for Research and Education. In the past, ADNI has also received funding from the National Institute of Biomedical Imaging and Bioengineering, the Canadian Institutes of Health Research, and private sector contributions through the Foundation for the National Institutes of Health (FNIH) including generous contributions from the following: AbbVie, Alzheimer’s Association; Alzheimer’s Drug Discovery Foundation; Araclon Biotech; BioClinica, Inc.; Biogen; Bristol-Myers Squibb Company; CereSpir, Inc.; Cogstate; Eisai Inc.; Elan Pharmaceuticals, Inc.; Eli Lilly and Company; EuroImmun; F. Hoffmann-La Roche Ltd and its affiliated company Genentech, Inc.; Fujirebio; GE Healthcare; IXICO Ltd.; Janssen Alzheimer Immunotherapy Research & Development, LLC.; Johnson & Johnson Pharmaceutical Research &Development LLC.; Lumosity; Lundbeck; Merck & Co., Inc.; Meso Scale Diagnostics, LLC.; NeuroRx Research; Neurotrack Technologies; Novartis Pharmaceuticals Corporation; Pfizer Inc.; Piramal Imaging; Servier; Takeda Pharmaceutical Company; and Transition Therapeutics. Data used in the preparation of this article were obtained on July 2, 2025, from the Parkinson’s Progression Markers Initiative (PPMI) database (www.ppmi-info.org/access-data-specimens/download-data; RRID: SCR_006431). For current information on the study, visit www.ppmi-info.org. PPMI – a public-private partnership – is funded by the Michael J. Fox Foundation for Parkinson’s Research and funding partners, including AbbVie, Alamar Biosciences, Aligning Science Across Parkinson’s (ASAP), Arrowhead Pharma, Arvinas, AskBio, BIAL, BioArctic, Biohaven, BlueRock Therapeutics, Bristol Myers Squibb, Calico Labs, Capsida Biotherapeutics, Critical Path Institute, DaCapo Brainscience, Denali, Edmond J. Safra Foundation, Eli Lilly, Gain Therapeutics, GE Healthcare, Genentech, GSK, Insitro, Johnson & Johnson Innovative Medicine, Lundbeck, Merck, Neumora, Neuron23, Novarti, Olink, Regeneron, Roche, Sanofi, Tenvie, UCB, Vanqua Bio, Voyager Therapeutics, The Weston Family Foundation. We thank María Renata Morales Miranda and Zyanya Galilea Estrada Meneses for their invaluable technical assistance with the systematic review. This work was derived from the thesis project of María Fernanda Zambrano-Astorga at the Posgrado en Ciencias de la Vida, CICESE, supported by a scholarship from SECIHTI (No. 4003993). The graphical abstract was created using biorender.com.

## Authors contributions

M.F.Z.A.: Conceptualization, data curation, research data, formal analysis, validation, visualization, writing, review & editing. A.M.U.: Conceptualization, data curation, formal analysis, funding acquisition, writing, review & editing. The manuscript’s contents have been approved by all the co-authors, and they have provided consent for its publication.

## Declaration of generative AI and AI-assisted technologies in the writing process

During the preparation of this work, the authors used ChatGPT/Perplexity and Copilot to assist with language refinement and to improve clarity of expression. After using these tools, the authors reviewed and edited the content as needed and takes full responsibility for the content of the published article.

## Methods

| REAGENT OR RESOURCE | SOURCE | IDENTIFIER |
| --- | --- | --- |
| Deposited data |  |  |
| ADNI Cruchaga lab CSF SOMAscan7k 20230620-2 | ADNI | https://adni.loni.usc.edu |
| PPMI Project 151 | PPMI<sup>38</sup> | https://www.ppmi-info.org/ |
| External studies | Bader et al.<sup>27</sup> | https://doi.org/10.15252/msb.20199356 |
|  | Bai et al.<sup>28</sup> | https://doi.org/10.1016/j.neuron.2019.12.015 |
|  | Higginbotham et al.<sup>4</sup> | https://doi.org/10.1126/sciadv.aaz9360 |
|  | Liu et al.<sup>35</sup> | https://doi.org/10.3390/ijms241814225 |
|  | Sathe et al.<sup>34</sup> | https://doi.org/10.1002/prca.201800105 |
|  | van der Ende et al.<sup>25</sup> | https://doi.org/10.1093/brain/awad213 |
|  | Wang et al.<sup>37</sup> | https://doi.org/10.1186/s13024-020-00384-6 |
|  | Bangs et al.<sup>21</sup> | https://doi.org/10.1101/2025.03.14.643332 |
|  | Johnson et al.<sup>12</sup> | https://doi.org/10.1038/s41591-020-0815-6 |
|  | Tao et al.<sup>36</sup> | https://doi.org/10.1016/j.xinn.2023.100544 |
| Software and algorithms |  |  |
| R 4.3.0 | Open source | https://cran.r-project.org/ |
| UniProt | Open source | https://www.uniprot.org/ |
| Seattle Alzheimer's Disease Cell Atlas | Michael Hawrylycz et al.<sup>99</sup> | https://doi.org/10.1038/s43587-024-00719-8 |
| SEA-AD Comparative Viewer | Gabitto et al.<sup>57</sup> | https://knowledge.brain-map.org/data/5IU4U8BP711TR6KZ843/2CD0HDC5PS6A58T0P6E/compare |
| NeuroPro | Askenazi et al.<sup>53</sup> | https://doi.org/10.1038/s41467-023-40208-x |
| Gene Ontology database | Aleksander et al.<sup>63</sup> | https://doi.org/10.1093/genetics/iyad031 |
| Reactome database | Gillespie et al.<sup>64</sup> | https://doi.org/10.1093/nar/gkab1028 |
| KEGG database | Kanehisa et al.<sup>100</sup> | https://doi.org/10.1093/nar/28.1.27 |
| pROC 1.18.5 | Robin et al.<sup>101</sup> | https://doi.org/10.1186/1471-2105-12-77 |
| ggplot2 3.3.3 | Wickham<sup>102</sup> | https://ggplot2.tidyverse.org |
| pheatmap 1.0.12 | Kolde<sup>103</sup> | https://github.com/raivokolde/pheatmap |
| Survival 3.7.0 | Therneau<sup>104</sup> | https://doi.org/10.32614/CRAN.package.survival |
| GOplot 1.0.2 | Walter et al.<sup>105</sup> | https://doi.org/10.1093/bioinformatics/btv300 |
| limma 3.62.2 | Ritchie et al.<sup>106</sup> | https://doi.org/10.1093/nar/gkv007 |
| Org.Hs.eg.db 3.22.0 | Carlson et al.<sup>107</sup> | https://doi.org/10.18129/B9.bioc.org.Hs.eg.db |
| clusterProfiler 4.0 | Wu et al.<sup>108</sup> | https://doi.org/10.1016/j.xinn.2021.100141 |
| Custom analysis codes | This paper | https://zenodo.org/records/19687369 |

### Experimental model and study participant details

#### Study design

This study aimed to identify a cerebrospinal fluid (CSF) proteomic signature for the diagnosis and prognosis of Alzheimer’s disease (AD) using a reproducibility-centered data mining framework; and to performed heat-to-head comparasion of this signature against previously published proteomic signatures. The study followed a four-step analytical process. First, a systematic literature review was conducted to identify published CSF proteomics studies eligible for the discovery and validation phases, based on predefined inclusion and exclusion criteria. Second, the discovery phase included eight independent cohorts (**Table 1**) comprising 759 participants, from which proteins showing consistent differential abundance between AD and control groups—defined as a log₂ fold change (FC) ± 0.6 and *p* < 0.05—were identified in at least two studies. These reproducible dysregulated proteins were retained as candidate biomarkers. Third, the candidate proteomic signature derived from the discovery phase was validated across three CSF proteomics cohorts (**Table 1**) encompassing 1,198 subjects. To evaluate disease specificity, we tested diagnostic metrics in a non-AD cognitively impaired subjects. Diagnostic performance was assessed by receiver operating characteristics (ROC) curve analysis, along with sensitivity and specificity estimations derived from optimal thresholds. For comparative evaluation, thirteen published CSF proteomic signatures were analyzed under the same parameters and cohorts. Prognostic performance—from cognitively uninpaired to A⁺T⁺ MCI and from A⁺T⁺ MCI to A⁺T⁺+dementia—was assessed using Cox proportional hazard models. Finally, molecular, cellular, and physiological insights into the signature proteins were examined through gene ontology (GO) and pathway enrichment analyses, integration with single-nucleus brain transcriptomics, and correlations with cognitive performance and neuroanatomical measures.

#### Proteomics datasets

##### Discovery studies

Eight published CSF proteomics studies were included in the discovery phase: Bader et al.^27^ (Maldenburg and Sweden cohorts), Bai et al.^28^, Higginbotham et al.^4^ (Discovery and Replication 4 cohorts), Liu et al.^35^ Sathe et al.^34^, Tao et al.^36^, van der Ende et al.^25^, and Wang et al.^37^, as summarized in **Table 1**. Comprehensive information for each dataset—including participant demographics, proteomic methodology, and data acquisition parameters—is provided in **Table S1**.

##### Validation studies and cohorts

Five independent cohorts were examined for the diagnosis performance, comprising two for model training and three independent cohorts for validation (**Table 1**). Training was performed using the Bader et al.^27^ (Sweden) and Tao et al.^36^ cohorts, while validation was conducted using ADNI, Bangs et al. (Emory Goizueta Alzheimer’s Disease Research Center [ADRC]) and Healthy Aging Study [EHBS])^21^, and Johnson et al. (Emory ADRC and EHBS)^12^ cohorts. In addition, the PPMI^38^ cohort was analyzed to evaluate diagnostic specificity in non-AD neurodegenerative conditions. Detailed descriptions of all validation datasets, including participant characteristics, proteomic and data processing methods, are presented in **Table S1**.

##### ADNI

CSF proteomics, clinical, and demographics data used in this study were obtained from the Alzheimer’s Disease Neuroimaging Initiative (ADNI) database (adni.loni.usc.edu). The ADNI was launched in 2003 as a public-private partnership, led by Principal Investigator Michael W. Weiner, MD. The primary goal of ADNI has been to test whether serial magnetic resonance imaging (MRI), positron emission tomography (PET), other biological markers, and clinical and neuropsychological assessment can be combined to measure the progression of mild cognitive impairment (MCI) and early Alzheimer’s disease (AD).

##### PPMI

The PPMI^38^, launched in 2010 by the Michael J. Fox Foundation, is a large-scale, longitudinal study aimed at identifying and validating biomarkers of Parkinson disease (PD). The study seeks to enroll approximately 4,000 participants for detailed clinical and imaging assessments and an additional 50,000 individuals for genetic screening and related PD evaluations. Participants undergo CSF collection, MRI scans, motor and cognitive assessments, and other tests to comprehensively characterize PD phenotypes. Further details on the PPMI cohort and study design are available at https://www.ppmi-info.org/.

### Method details

#### Literature review and data selection

A systematic literature review was conducted to identify CSF proteomics studies that included both AD and control participants. Searches were performed in PubMed using combinations of keywords and Boolean operators across all fields (“Alzheimer’s disease” AND “cerebrospinal fluid” AND “proteomics”), (“Alzheimer’s disease” AND “CSF” AND “proteomics”), (“Alzheimer’s disease” AND “cerebrospinal fluid” AND “biomarkers”), (“Alzheimer’s disease” AND “CSF” AND “biomarkers”), (“Alzheimer’s disease” AND “cerebrospinal fluid” AND “mass spectrometry”), and (“Alzheimer’s disease” AND “CSF” AND “mass spectrometry”). Articles published up to November 11, 2024 (date of search) were considered eligible. Studies were manually screened and included if they met the following criteria: (1) original research reporting either a quantitative protein abundance matrix or a list of differentially abundant proteins (DAPs) with corresponding *p*-values and FC; (2) conducted in human subjects; (3) comparing AD-diagnosed individuals —confirmed by CSF or imaging biomarkers (A⁺T⁺ or A⁺) and standardized cognitive assessments consistent with the National Institute on Aging and Alzheimer’s Association (NIA-AA)^1^, the National Institute of Neurological and Communicative Disorders and Stroke and the Alzheimer’s Disease and

Related Disorders Association (NINCDS-ADRDA)^109^ or the International Working Group ^2^ diagnostic criteria or Consortium to Establish a Registry for Alzheimer’s Disease (CERAD)^110^ and Braak^111^ for post-mortem samples— with healthy controls (A^-^T^-^, and CU); (4) included at least three subjects per group; (5) employed untargeted proteomic approaches; and (6) used high-throughput data acquisition platforms such as mass spectrometry, SomaScan, or Olink, with clear descriptions of acquisition techniques and processing methods. Studies were excluded if they (1) lacked complete sample handling details; (2) proteomics data derived from animal or *in vitro* models; (3) used non-standard diagnostic procedures; (4) included fewer than three subjects per group; (5) contained missing or incomplete datasets; (6) employed targeted proteomic methods; (7) inferred proteomic changes from GWAS or transcriptomic data; and (8) used low-throughput or semi-quantitative techniques such as Selected Reaction Monitoring (SRM), Multiple Reaction Monitoring (MRM), Parallel Reaction Monitoring (PRM), Western blotting, or two-dimensional gel electrophoresis.

#### Data extraction and processing

For the discovery phase, relevant metadata and proteomic results were extracted from each included study and compiled into a local database (**Table S1**). Only identified DAPs were retained, defined by a significance threshold of *p* < 0.05 (Student’s t-test or ANOVA) and a log₂ FC ± 0.6. For independent validation, quantitative protein abundance matrices from Bangs et al.^21^ (Synapse ID: syn65461849) and Johnson et al.^12^ (Synapse ID: syn20821165) were downloaded from https://www.synapse.org/ after registration and approval. We used the preprocessed data provided by the original authors, which included batch correction, normalization, and log₂ transformation. Datasets from Tao et al.^36^ and Bader et al.^27^ were obtained from the supplementary materials of the respective publications and processed using MetaboAnalyst v6.0^112^ (https://www.metaboanalyst.ca/). Data processing included batch correction, exclusion of proteins with > 80% missing values, quantile normalization, and log₂ transformation. Harmonization across cohort matrices was achieved through z-score transformation. For ADNI and PPMI, demographic data, unregressed proteomic matrices, and analyte metadata were directly downloaded from adni.loni.usc.edu and ppmi-info.org repositories. For both datasets, analyses were performed using the authors’ normalized and log₂-transformed proteomic matrices.

#### AD classification

Detailed information on AD classification for each included study is provided in **Table 1**. For the discovery phase, we utilized the original authors classification, which followed either the NIA-AA^1^ or IWG^2^ clinical guidelines. For biological classification, AT biomarkers were assessed using CSF measures of t-Tau/Aβ_42_, p-Tau/Aβ_42_, Aβ_42_/Aβ_40_, and Aβ_42_, as well as PET ^18^F-AV-45 imaging. In postmortem cases, classification was based on Braak staging^111^ and the CERAD^110^ neuropathological criteria. For clinical classification, in addition to the IWG framework, diagnostic evaluation followed the criteria of the NINCDS-ADRDA^109^ and thr Diagnostic and Statistical Manual of Mental Disorders (DSM). These assessments incorporated the CERAD neuropsychological test battery, standard cognitive and functional measures, and comprehensive medical evaluations. For the validation phase, we stratified subjects from the ADNI and Bangs et al.^21^ cohorts consisted of biological and combined biological–clinical diagnosis according to NIA-AA)^1^ and IWG^2^ clinical guidelines. For ADNI cohort biological classification was based on p-Tau/Aβ_42_ ratio and clinical classification was stablish by authors based on clinical and functional assessment. For Bangs et al.^21^ cohort biological classification was based on t-Tau/Aβ_42_ ratio and clinical classification by the Montreal Cognitive Assesment (MoCA).

### Quantification and statistical analysis

#### Biomarker discovery

The initial biomarker panel was obtained by selecting DAPs—defined as *p* < 0.05 and a log₂ FC ± 0.6—in at least two independent CSF proteomics studies, with consistent directionality of change (either up- or downregulation). Protein co-occurrence across studies was assessed using UpSet plots, while the directionality and magnitude of abundance changes were visualized as heatmaps generated in RStudio using the pheatmap package (v1.0.12) (https://github.com/raivokolde/pheatmap)^103^. Among proteins meeting reproducibility criteria, only those detected across the validation cohorts were retained as biomarker candidates. These candidates were subsequently evaluated in multivariable diagnostic models, and the final size of the biomarker panel was determined based on model performance.

#### Diagnostic performance

The diagnostic performance of the proposed signature and previously published CSF proteomic signatures, used for comparative purposes, was evaluated using ROC curve analysis implemented in RStudio (v4.4.3) with the pROC package (v1.18.5)^101^ across four validation cohorts ADNI (n = 702), Bangs et al.^21^ (n = 431), Johnson et al.^12^ (n = 65), and PPMI^38^ (n = 798).

Diagnostic classification was for both biological (A⁺T⁺ vs. A⁻T⁻ and A⁺ vs. A⁻) and combined biological–clinical comparisons (A⁺T⁺ CI [MCI or dementia] vs. A⁻T⁻ CU) for AD diagnosis. For selectivity analysis, comparing individuals with cognitive impairment due to non-AD causes (A⁻T⁻) and PD against healthy controls. Due to the nature differences of proteomic platforms used for data acquisition, we performed two different validation models, one for SomaScan and the other for liquid chromatography coupled to mass spectrometry (LC-MS/MS) derived data. For SomaScan data (ADNI and PPMI), a five-fold cross-validation logistic regression model with ten repetitions was implemented. For LC-MS/MS data, logistic regression models were trained using Bader et al.^27^ and Tao et al.^36^ cohorts and validated independently in Bangs et al.^21^ and Johnson et al.^12^. Visualization of results was performed with ggplot2 (v3.3.3) (https://ggplot2.tidyverse.org), and sensitivity and specificity were determined using Youden’s J statistic to identify optimal cut-off thresholds. To assess disease selectivity, the signature was further tested among subjects with PD (PD vs controls, n = 798) and in subsets of individuals with CI not linked to AD (A⁻T⁻ MCI vs A⁻T⁻ CU n = 488). Finally, to contextualize diagnostic performance, the proposed signature was benchmarked against thirteen previously published CSF-based proteomic signatures. Relevant studies were identified through a systematic PubMed search as previously described. Comparative ROC analyses were performed using the same methodological framework, with minor adaptations to enable direct comparison.

#### Prognosis performance

To assess the prognostic capacity of PPAV11 and comparative signatures time-to-event analyses were performed using Cox proportional hazard models implemented with the survival package (v3.7.0) (https://CRAN.R-project.org/package=survival)^104^ in RStudio (v4.4.3). The ADNI cohort was selected for this analysis as it was the only dataset with longitudinal follow-up data. Participants with at least two visits over a follow-up period ranging from six months to 12.5 years were included. Disease progression was modeled across two transitions: from CU to A⁺T⁺ MCI (n = 141), and from A⁺T⁺ MCI to A⁺T⁺ dementia (n = 174). Participants were stratified into high- and low-scores (based on median protein level values), and survival functions were compared between strata. Hazzard ratios comparassions were visualized in forest plots and progression trajectories were visualized using Kaplan–Meier curves. Statistical significance (*p* < 0.05) was determined using the Wald test. All graphical outputs were generated with ggplot2 (v3.3.3) (https://ggplot2.tidyverse.org)^102^.

#### Correlation analysis with clinical variables

Associations between PPAV11 protein levels and clinical variables were assessed using correlation analysis implemented in R (v4.4.3) with the limma package (v3.62.2)^106^. Clinical variables encompassed cognitive and functional assessments, including MoCA, Mini-Mental State Examination (MMSE), Alzheimer’s Disease Assessment Scale (ADAS13 and ADASQ4), Clinical Dementia Rating (CDR), Functional Activities Questionnaire (FAQ), Rey Auditory Verbal Learning Test (RAVLT), and Logical Memory Delayed Recall (LDELTOTAL). Additionally, structural MRI-derived volumetric measures were incorporated, including total brain volume, intracranial volume, ventricular volume, and regional volumes of the hippocampus, middle temporal lobe, entorhinal cortex, and fusiform gyrus. Sex and age were included as covariates in all models. Results were visualized using ggplot2 (v3.3.3) (https://ggplot2.tidyverse.org)^102^. Statical analysis was performed in the limma analysis and corrected with Benjamini-Hochberg analysis. Significance of correlations was defined as adjusted p < 0.05.

#### Functional enrichment analysis

Functional enrichment analysis was conducted in RStudio (v4.4.3) using org.Hs.eg.db package (v3.22.0)^107^ along with biological annotation databases to identify overrepresented biological processes, pathways, and molecular functions associated with the PPAV11 proteins. Analyses incorporated the GO^63^, REACTOME^64^ (via the ReactomePA package, v1.50.0), and KEGG^100^ databases. Visualization of enriched terms and pathway relationships was performed using ggplot2 (v3.3.3) (https://ggplot2.tidyverse.org)^102^ and circlize (v0.4.16)^113^. Terms were considered significantly enriched when passing the false discovery rate threshold (FDR < 0.05).

#### Regional Brain Dysregulation Analysis

To examine the regional dysregulation of PPAV11 proteins across the brain, we utilized the NeuroPro database^53^ which integrates data from 32 large-scale proteomic studies encompassing 13 brain regions from individuals with and without AD. Each PPAV11 protein was individually queried in the database, and brain regions where protein abundance exhibited a FC ≥ 1.5 or ≤ 0.667 with FDR < 0.05 or adjusted-p < 0.05 in AD vs controls comparisons were retained. A brain map was then manually constructed to visualize the regions and corresponding PPAV11 proteins meeting the criteria for differential abundance.

#### Brain cell-type enrichment analysis

Cell-type enrichment was assessed using the Transcriptomics Comparative Viewer available through the Brain Knowledge Platform (https://brain-map.org/bkp) from the Allen Institute. To characterize the brain cell subtypes in which PPAV11 transcripts are expressed, we analyzed single-nucleus RNA sequencing data obtained from the middle temporal gyrus (MTG) of five neurotypical human donors^57^. Cells were stratified into major populations, including GABAergic and glutamatergic neurons, as well as non-neuronal cell types. Additionally, we examined the pseudoprogression of PPAV11 expression in the MTG using the 10x Seattle Alzheimer’s Disease (SEA-AD) dataset^114^, which comprises single-nucleus transcriptomic profiles from 84 aged brain donors stratified by pseudoprogression score (CPS)^57,99^.

## References

1. Jack, C.R., Jr., Andrews, J.S., Beach, T.G., Buracchio, T., Dunn, B., Graf, A., Hansson, O., Ho, C., Jagust, W., McDade, E., et al. (2024). Revised criteria for diagnosis and staging of Alzheimer’s disease: Alzheimer’s Association Workgroup. Alzheimers Dement 20, 5143–5169. 10.1002/alz.13859.

2. Dubois, B., Villain, N., Schneider, L., Fox, N., Campbell, N., Galasko, D., Kivipelto, M., Jessen, F., Hanseeuw, B., Boada, M., et al. (2024). Alzheimer Disease as a Clinical-Biological Construct-An International Working Group Recommendation. JAMA Neurol 81, 1304–1311. 10.1001/jamaneurol.2024.3770.

3. Oh, H.S., Urey, D.Y., Karlsson, L., Zhu, Z., Shen, Y., Farinas, A., Timsina, J., Duggan, M.R., Chen, J., Guldner, I.H., et al. (2025). A cerebrospinal fluid synaptic protein biomarker for prediction of cognitive resilience versus decline in Alzheimer’s disease. Nat Med 31, 1592–1603. 10.1038/s41591-025-03565-2.

4. Higginbotham, L., Ping, L., Dammer, E.B., Duong, D.M., Zhou, M., Gearing, M., Hurst, C., Glass, J.D., Factor, S.A., Johnson, E.C.B., et al. (2020). Integrated proteomics reveals brain-based cerebrospinal fluid biomarkers in asymptomatic and symptomatic Alzheimer’s disease. Sci Adv 6. 10.1126/sciadv.aaz9360.

5. Park, S.A., Han, S.M., and Kim, C.E. (2020). New fluid biomarkers tracking non-amyloid-beta and non-tau pathology in Alzheimer’s disease. Exp Mol Med 52, 556–568. 10.1038/s12276-020-0418-9.

6. Johnson, E.C.B., Bian, S., Haque, R.U., Carter, E.K., Watson, C.M., Gordon, B.A., Ping, L., Duong, D.M., Epstein, M.P., McDade, E., et al. (2023). Cerebrospinal fluid proteomics define the natural history of autosomal dominant Alzheimer’s disease. Nat Med 29, 1979–1988. 10.1038/s41591-023-02476-4.

7. Rayaprolu, S., Higginbotham, L., Bagchi, P., Watson, C.M., Zhang, T., Levey, A.I., Rangaraju, S., and Seyfried, N.T. (2021). Systems-based proteomics to resolve the biology of Alzheimer’s disease beyond amyloid and tau. Neuropsychopharmacology 46, 98–115. 10.1038/s41386-020-00840-3.

8. Tijms, B.M., Vromen, E.M., Mjaavatten, O., Holstege, H., Reus, L.M., van der Lee, S., Wesenhagen, K.E.J., Lorenzini, L., Vermunt, L., Venkatraghavan, V., et al. (2024). Cerebrospinal fluid proteomics in patients with Alzheimer’s disease reveals five molecular subtypes with distinct genetic risk profiles. Nat Aging 4, 33–47. 10.1038/s43587-023-00550-7.

9. Young-Pearse, T.L., Lee, H., Hsieh, Y.C., Chou, V., and Selkoe, D.J. (2023). Moving beyond amyloid and tau to capture the biological heterogeneity of Alzheimer’s disease. Trends Neurosci 46, 426–444. 10.1016/j.tins.2023.03.005.

10. Jansen, I.E., van der Lee, S.J., Gomez-Fonseca, D., de Rojas, I., Dalmasso, M.C., Grenier-Boley, B., Zettergren, A., Mishra, A., Ali, M., Andrade, V., et al. (2022). Genome-wide meta-analysis for Alzheimer’s disease cerebrospinal fluid biomarkers. Acta Neuropathol 144, 821–842. 10.1007/s00401-022-02454-z.

11. Bellenguez, C., Kucukali, F., Jansen, I.E., Kleineidam, L., Moreno-Grau, S., Amin, N., Naj, A.C., Campos-Martin, R., Grenier-Boley, B., Andrade, V., et al. (2022). New insights into the genetic etiology of Alzheimer’s disease and related dementias. Nat Genet 54, 412–436. 10.1038/s41588-022-01024-z.

12. Johnson, E.C.B., Dammer, E.B., Duong, D.M., Ping, L., Zhou, M., Yin, L., Higginbotham, L.A., Guajardo, A., White, B., Troncoso, J.C., et al. (2020). Large-scale proteomic analysis of Alzheimer’s disease brain and cerebrospinal fluid reveals early changes in energy metabolism associated with microglia and astrocyte activation. Nat Med 26, 769–780. 10.1038/s41591-020-0815-6.

13. Kunkle, B.W., Grenier-Boley, B., Sims, R., Bis, J.C., Damotte, V., Naj, A.C., Boland, A., Vronskaya, M., van der Lee, S.J., Amlie-Wolf, A., et al. (2019). Genetic meta-analysis of diagnosed Alzheimer’s disease identifies new risk loci and implicates Abeta, tau, immunity and lipid processing. Nat Genet 51, 414–430. 10.1038/s41588-019-0358-2.

14. Rathore, S., Dammer, E.B., Shantaraman, A., Wu, F., Duong, D.M., Fox, E.J., Johnson, E.C.B., Lah, J.J., Alzheimer’s Disease Neuroimaging, I., Seyfried, N.T., and Levey, A.I. (2025). CSF Proteomics and Machine Learning Reveal Distinct Stages Across the Alzheimer’s Disease Continuum. medRxiv. 10.1101/2025.11.05.25339138.

15. Imam, F., Saloner, R., Vogel, J.W., Krish, V., Abdel-Azim, G., Ali, M., An, L., Anastasi, F., Bennett, D., Pichet Binette, A., et al. (2025). The Global Neurodegeneration Proteomics Consortium: biomarker and drug target discovery for common neurodegenerative diseases and aging. Nat Med 31, 2556–2566. 10.1038/s41591-025-03834-0.

16. Hansson, O. (2021). Biomarkers for neurodegenerative diseases. Nat Med 27, 954–963. 10.1038/s41591-021-01382-x.

17. Halama, A., Zaghlool, S., Thareja, G., Kader, S., Al Muftah, W., Mook-Kanamori, M., Sarwath, H., Mohamoud, Y.A., Stephan, N., Ameling, S., et al. (2024). A roadmap to the molecular human linking multiomics with population traits and diabetes subtypes. Nat Commun 15, 7111. 10.1038/s41467-024-51134-x.

18. Heo, G., Xu, Y., Wang, E., Ali, M., Oh, H.S., Moran-Losada, P., Anastasi, F., Gonzalez Escalante, A., Puerta, R., Song, S., et al. (2025). Large-scale plasma proteomic profiling unveils diagnostic biomarkers and pathways for Alzheimer’s disease. Nat Aging 5, 1114–1131. 10.1038/s43587-025-00872-8.

19. Ali, M., Timsina, J., Western, D., Liu, M., Beric, A., Budde, J., Do, A., Heo, G., Wang, L., Gentsch, J., et al. (2025). Multi-cohort cerebrospinal fluid proteomics identifies robust molecular signatures across the Alzheimer disease continuum. Neuron 113, 1363–1379 e1369. 10.1016/j.neuron.2025.02.014.

20. Hablitz, L.M., and Nedergaard, M. (2025). Cerebrospinal fluid flow modulates brain health. J Clin Invest 135. 10.1172/JCI197202.

21. Bangs, M.C., Gadhavi, J., Carter, E.K., Ping, L., Duong, D.M., Dammer, E.B., Wu, F., Shantaraman, A., Fox, E.J., Johnson, E.C.B., et al. (2025). Proteomic Subtyping of Alzheimer’s Disease CSF links Blood-Brain Barrier Dysfunction to Reduced levels of Tau and Synaptic Biomarkers. bioRxiv. 10.1101/2025.03.14.643332.

22. Guo, Y., Chen, S.D., You, J., Huang, S.Y., Chen, Y.L., Zhang, Y., Wang, L.B., He, X.Y., Deng, Y.T., Zhang, Y.R., et al. (2024). Multiplex cerebrospinal fluid proteomics identifies biomarkers for diagnosis and prediction of Alzheimer’s disease. Nat Hum Behav 8, 2047–2066. 10.1038/s41562-024-01924-6.

23. Modeste, E.S., Ping, L., Watson, C.M., Duong, D.M., Dammer, E.B., Johnson, E.C.B., Roberts, B.R., Lah, J.J., Levey, A.I., and Seyfried, N.T. (2023). Quantitative proteomics of cerebrospinal fluid from African Americans and Caucasians reveals shared and divergent changes in Alzheimer’s disease. Mol Neurodegener 18, 48. 10.1186/s13024-023-00638-z.

24. Sung, Y.J., Yang, C., Norton, J., Johnson, M., Fagan, A., Bateman, R.J., Perrin, R.J., Morris, J.C., Farlow, M.R., Chhatwal, J.P., et al. (2023). Proteomics of brain, CSF, and plasma identifies molecular signatures for distinguishing sporadic and genetic Alzheimer’s disease. Sci Transl Med 15, eabq5923. 10.1126/scitranslmed.abq5923.

25. van der Ende, E.L., In ’t Veld, S., Hanskamp, I., van der Lee, S., Dijkstra, J.I.R., Hok, A.H.Y.S., Blujdea, E.R., van Swieten, J.C., Irwin, D.J., Chen-Plotkin, A., et al. (2023). CSF proteomics in autosomal dominant Alzheimer’s disease highlights parallels with sporadic disease. Brain 146, 4495–4507. 10.1093/brain/awad213.

26. Del Campo, M., Peeters, C.F.W., Johnson, E.C.B., Vermunt, L., Hok, A.H.Y.S., van Nee, M., Chen-Plotkin, A., Irwin, D.J., Hu, W.T., Lah, J.J., et al. (2022). CSF proteome profiling across the Alzheimer’s disease spectrum reflects the multifactorial nature of the disease and identifies specific biomarker panels. Nat Aging 2, 1040–1053. 10.1038/s43587-022-00300-1.

27. Bader, J.M., Geyer, P.E., Muller, J.B., Strauss, M.T., Koch, M., Leypoldt, F., Koertvelyessy, P., Bittner, D., Schipke, C.G., Incesoy, E.I., et al. (2020). Proteome profiling in cerebrospinal fluid reveals novel biomarkers of Alzheimer’s disease. Mol Syst Biol 16, e9356. 10.15252/msb.20199356.

28. Bai, B., Wang, X., Li, Y., Chen, P.C., Yu, K., Dey, K.K., Yarbro, J.M., Han, X., Lutz, B.M., Rao, S., et al. (2020). Deep Multilayer Brain Proteomics Identifies Molecular Networks in Alzheimer’s Disease Progression. Neuron 105, 975–991 e977. 10.1016/j.neuron.2019.12.015.

29. van Zalm, P.W., Ahmed, S., Fatou, B., Schreiber, R., Barnaby, O., Boxer, A., Zetterberg, H., Steen, J.A., and Steen, H. (2023). Meta-analysis of published cerebrospinal fluid proteomics data identifies and validates metabolic enzyme panel as Alzheimer’s disease biomarkers. Cell Rep Med 4, 101005. 10.1016/j.xcrm.2023.101005.

30. Querol-Vilaseca, M., Colom-Cadena, M., Pegueroles, J., San Martin-Paniello, C., Clarimon, J., Belbin, O., Fortea, J., and Lleo, A. (2017). YKL-40 (Chitinase 3-like I) is expressed in a subset of astrocytes in Alzheimer’s disease and other tauopathies. J Neuroinflammation 14, 118. 10.1186/s12974-017-0893-7.

31. Zhang, Q., Wang, S.S., Zhang, Z., and Chu, S.F. (2025). PKM2-mediated metabolic reprogramming of microglia in neuroinflammation. Cell Death Discov 11, 149. 10.1038/s41420-025-02453-5.

32. Balcomb, K., Johnston, C., Kavanagh, T., Leitner, D., Schneider, J., Halliday, G., Wisniewski, T., Sunde, M., and Drummond, E. (2024). SMOC1 colocalizes with Alzheimer’s disease neuropathology and delays Abeta aggregation. Acta Neuropathol 148, 72. 10.1007/s00401-024-02819-6.

33. Oosthoek, M., Vijverberg, E.G.B., Blujdea, E.R., Veld, S., Aviles, M.P., Zsadanyi, S.E., Hok, A.H.Y.S., Visser, A., van der Flier, W.M., Barkhof, F., et al. (2025). CHIT1 and DDAH1 levels relate to amyloid-related imaging abnormalities risk profile in Alzheimer’s disease patients. Alzheimers Res Ther 17, 169. 10.1186/s13195-025-01799-3.

34. Sathe, G., Na, C.H., Renuse, S., Madugundu, A.K., Albert, M., Moghekar, A., and Pandey, A. (2019). Quantitative Proteomic Profiling of Cerebrospinal Fluid to Identify Candidate Biomarkers for Alzheimer’s Disease. Proteomics Clin Appl 13, e1800105. 10.1002/prca.201800105.

35. Liu, P., Li, L., He, F., Meng, F., Liu, X., Su, Y., Su, X., Luo, B., and Peng, G. (2023). Identification of Candidate Biomarkers of Alzheimer’s Disease via Multiplex Cerebrospinal Fluid and Serum Proteomics. Int J Mol Sci 24. 10.3390/ijms241814225.

36. Tao, Q.Q., Cai, X., Xue, Y.Y., Ge, W., Yue, L., Li, X.Y., Lin, R.R., Peng, G.P., Jiang, W., Li, S., et al. (2024). Alzheimer’s disease early diagnostic and staging biomarkers revealed by large-scale cerebrospinal fluid and serum proteomic profiling. Innovation (Camb) 5, 100544. 10.1016/j.xinn.2023.100544.

37. Wang, H., Dey, K.K., Chen, P.C., Li, Y., Niu, M., Cho, J.H., Wang, X., Bai, B., Jiao, Y., Chepyala, S.R., et al. (2020). Integrated analysis of ultra-deep proteomes in cortex, cerebrospinal fluid and serum reveals a mitochondrial signature in Alzheimer’s disease. Mol Neurodegener 15, 43. 10.1186/s13024-020-00384-6.

38. Marek, K., Chowdhury, S., Siderowf, A., Lasch, S., Coffey, C.S., Caspell-Garcia, C., Simuni, T., Jennings, D., Tanner, C.M., Trojanowski, J.Q., et al. (2018). The Parkinson’s progression markers initiative (PPMI) - establishing a PD biomarker cohort. Ann Clin Transl Neurol 5, 1460–1477. 10.1002/acn3.644.

39. Hou, X., Qiu, Y., Li, H., Yan, Y., Zhao, D., Ji, S., Ni, J., Zhang, J., Liu, K., Qing, H., and Quan, Z. (2025). Machine-learning based strategy identifies a robust protein biomarker panel for Alzheimer’s disease in cerebrospinal fluid. Alzheimers Res Ther 17, 147. 10.1186/s13195-025-01789-5.

40. Shen, Y., Timsina, J., Heo, G., Beric, A., Ali, M., Wang, C., Yang, C., Wang, Y., Western, D., Liu, M., et al. (2024). CSF proteomics identifies early changes in autosomal dominant Alzheimer’s disease. Cell 187, 6309–6326 e6315. 10.1016/j.cell.2024.08.049.

41. Fagan, A.M., Roe, C.M., Xiong, C., Mintun, M.A., Morris, J.C., and Holtzman, D.M. (2007). Cerebrospinal fluid tau/beta-amyloid(42) ratio as a prediction of cognitive decline in nondemented older adults. Arch Neurol 64, 343–349. 10.1001/archneur.64.3.noc60123.

42. Salvado, G., Larsson, V., Cody, K.A., Cullen, N.C., Jonaitis, E.M., Stomrud, E., Kollmorgen, G., Wild, N., Palmqvist, S., Janelidze, S., et al. (2023). Optimal combinations of CSF biomarkers for predicting cognitive decline and clinical conversion in cognitively unimpaired participants and mild cognitive impairment patients: A multi-cohort study. Alzheimers Dement 19, 2943–2955. 10.1002/alz.12907.

43. Shiino, A., Shirakashi, Y., Ishida, M., Tanigaki, K., and Japanese Alzheimer’s Disease Neuroimaging, I. (2021). Machine learning of brain structural biomarkers for Alzheimer’s disease (AD) diagnosis, prediction of disease progression, and amyloid beta deposition in the Japanese population. Alzheimers Dement (Amst) 13, e12246. 10.1002/dad2.12246.

44. Li, G., Sokal, I., Quinn, J.F., Leverenz, J.B., Brodey, M., Schellenberg, G.D., Kaye, J.A., Raskind, M.A., Zhang, J., Peskind, E.R., and Montine, T.J. (2007). CSF tau/Abeta42 ratio for increased risk of mild cognitive impairment: a follow-up study. Neurology 69, 631–639. 10.1212/01.wnl.0000267428.62582.aa.

45. Galasko, D., Xiao, M., Xu, D., Smirnov, D., Salmon, D.P., Dewit, N., Vanbrabant, J., Jacobs, D., Vanderstichele, H., Vanmechelen, E., et al. (2019). Synaptic biomarkers in CSF aid in diagnosis, correlate with cognition and predict progression in MCI and Alzheimer’s disease. Alzheimers Dement (N Y) 5, 871–882. 10.1016/j.trci.2019.11.002.

46. Xiao, M.F., Xu, D., Craig, M.T., Pelkey, K.A., Chien, C.C., Shi, Y., Zhang, J., Resnick, S., Pletnikova, O., Salmon, D., et al. (2017). NPTX2 and cognitive dysfunction in Alzheimer’s Disease. Elife 6. 10.7554/eLife.23798.

47. Swanson, A., Willette, A.A., and Alzheimer’s Disease Neuroimaging, I. (2016). Neuronal Pentraxin 2 predicts medial temporal atrophy and memory decline across the Alzheimer’s disease spectrum. Brain Behav Immun 58, 201–208. 10.1016/j.bbi.2016.07.148.

48. Sauer, M., Gomes, B.F., Shahrouki, P., Lantero-Rodriguez, J., Montoliu-Gaya, L., Camporesi, E., Belbin, O., Alcolea, D., Kumar, A., Sharma, M., et al. (2025). Development and validation of a novel Simoa assay for NPTX2 in Alzheimer’s disease and Down syndrome. Alzheimers Dement 21, e70241. 10.1002/alz.70241.

49. Zhou, J., Wade, S.D., Graykowski, D., Xiao, M.F., Zhao, B., Giannini, L.A.A., Hanson, J.E., van Swieten, J.C., Sheng, M., Worley, P.F., and Dejanovic, B. (2023). The neuronal pentraxin Nptx2 regulates complement activity and restrains microglia-mediated synapse loss in neurodegeneration. Sci Transl Med 15, eadf0141. 10.1126/scitranslmed.adf0141.

50. Pini, L., Pievani, M., Bocchetta, M., Altomare, D., Bosco, P., Cavedo, E., Galluzzi, S., Marizzoni, M., and Frisoni, G.B. (2016). Brain atrophy in Alzheimer’s Disease and aging. Ageing Res Rev 30, 25–48. 10.1016/j.arr.2016.01.002.

51. Apostolova, L.G., and Cummings, J.L. (2008). Neuropsychiatric manifestations in mild cognitive impairment: a systematic review of the literature. Dement Geriatr Cogn Disord 25, 115–126. 10.1159/000112509.

52. Lerch, J.P., and Evans, A.C. (2005). Cortical thickness analysis examined through power analysis and a population simulation. Neuroimage 24, 163–173. 10.1016/j.neuroimage.2004.07.045.

53. Askenazi, M., Kavanagh, T., Pires, G., Ueberheide, B., Wisniewski, T., and Drummond, E. (2023). Compilation of reported protein changes in the brain in Alzheimer’s disease. Nat Commun 14, 4466. 10.1038/s41467-023-40208-x.

54. Salta, E., Lazarov, O., Fitzsimons, C.P., Tanzi, R., Lucassen, P.J., and Choi, S.H. (2023). Adult hippocampal neurogenesis in Alzheimer’s disease: A roadmap to clinical relevance. Cell Stem Cell 30, 120–136. 10.1016/j.stem.2023.01.002.

55. Palmqvist, S., Scholl, M., Strandberg, O., Mattsson, N., Stomrud, E., Zetterberg, H., Blennow, K., Landau, S., Jagust, W., and Hansson, O. (2017). Earliest accumulation of beta-amyloid occurs within the default-mode network and concurrently affects brain connectivity. Nat Commun 8, 1214. 10.1038/s41467-017-01150-x.

56. Johnson, J.K., Head, E., Kim, R., Starr, A., and Cotman, C.W. (1999). Clinical and pathological evidence for a frontal variant of Alzheimer disease. Arch Neurol 56, 1233–1239. 10.1001/archneur.56.10.1233.

57. Gabitto, M.I., Travaglini, K.J., Rachleff, V.M., Kaplan, E.S., Long, B., Ariza, J., Ding, Y., Mahoney, J.T., Dee, N., Goldy, J., et al. (2024). Integrated multimodal cell atlas of Alzheimer’s disease. Nat Neurosci 27, 2366–2383. 10.1038/s41593-024-01774-5.

58. Lao, Y., Xiao, M.F., Ji, S., Piras, I.S., Bonfitto, A., Song, S., Aldabergenova, A., Na, C.H., Sloan, J., Trejo, A., et al. (2025). Transcriptomic and protein analysis of human cortex reveals genes and pathways linked to NPTX2 disruption in Alzheimer’s disease. bioRxiv. 10.1101/2025.10.17.683150.

59. Sha, X., Lin, J., Wu, K., Lu, J., and Yu, Z. (2025). The TRPV1-PKM2-SREBP1 axis maintains microglial lipid homeostasis in Alzheimer’s disease. Cell Death Dis 16, 14. 10.1038/s41419-024-07328-8.

60. Lananna, B.V., McKee, C.A., King, M.W., Del-Aguila, J.L., Dimitry, J.M., Farias, F.H.G., Nadarajah, C.J., Xiong, D.D., Guo, C., Cammack, A.J., et al. (2020). Chi3l1/YKL-40 is controlled by the astrocyte circadian clock and regulates neuroinflammation and Alzheimer’s disease pathogenesis. Sci Transl Med 12. 10.1126/scitranslmed.aax3519.

61. Antonell, A., Mansilla, A., Rami, L., Llado, A., Iranzo, A., Olives, J., Balasa, M., Sanchez-Valle, R., and Molinuevo, J.L. (2014). Cerebrospinal fluid level of YKL-40 protein in preclinical and prodromal Alzheimer’s disease. J Alzheimers Dis 42, 901–908. 10.3233/JAD-140624.

62. Craig-Schapiro, R., Perrin, R.J., Roe, C.M., Xiong, C., Carter, D., Cairns, N.J., Mintun, M.A., Peskind, E.R., Li, G., Galasko, D.R., et al. (2010). YKL-40: a novel prognostic fluid biomarker for preclinical Alzheimer’s disease. Biol Psychiatry 68, 903–912. 10.1016/j.biopsych.2010.08.025.

63. Gene Ontology, C., Aleksander, S.A., Balhoff, J., Carbon, S., Cherry, J.M., Drabkin, H.J., Ebert, D., Feuermann, M., Gaudet, P., Harris, N.L., et al. (2023). The Gene Ontology knowledgebase in 2023. Genetics 224. 10.1093/genetics/iyad031.

64. Gillespie, M., Jassal, B., Stephan, R., Milacic, M., Rothfels, K., Senff-Ribeiro, A., Griss, J., Sevilla, C., Matthews, L., Gong, C., et al. (2022). The reactome pathway knowledgebase 2022. Nucleic Acids Res 50, D687–D692. 10.1093/nar/gkab1028.

65. Panyard, D.J., McKetney, J., Deming, Y.K., Morrow, A.R., Ennis, G.E., Jonaitis, E.M., Van Hulle, C.A., Yang, C., Sung, Y.J., Ali, M., et al. (2023). Large-scale proteome and metabolome analysis of CSF implicates altered glucose and carbon metabolism and succinylcarnitine in Alzheimer’s disease. Alzheimers Dement 19, 5447–5470. 10.1002/alz.13130.

66. Johnson, E.C.B., Carter, E.K., Dammer, E.B., Duong, D.M., Gerasimov, E.S., Liu, Y., Liu, J., Betarbet, R., Ping, L., Yin, L., et al. (2022). Large-scale deep multi-layer analysis of Alzheimer’s disease brain reveals strong proteomic disease-related changes not observed at the RNA level. Nat Neurosci 25, 213–225. 10.1038/s41593-021-00999-y.

67. Tijms, B.M., Gobom, J., Reus, L., Jansen, I., Hong, S., Dobricic, V., Kilpert, F., Ten Kate, M., Barkhof, F., Tsolaki, M., et al. (2020). Pathophysiological subtypes of Alzheimer’s disease based on cerebrospinal fluid proteomics. Brain 143, 3776–3792. 10.1093/brain/awaa325.

68. Heppner, F.L., Ransohoff, R.M., and Becher, B. (2015). Immune attack: the role of inflammation in Alzheimer disease. Nat Rev Neurosci 16, 358–372. 10.1038/nrn3880.

69. Butterfield, D.A., and Halliwell, B. (2019). Oxidative stress, dysfunctional glucose metabolism and Alzheimer disease. Nat Rev Neurosci 20, 148–160. 10.1038/s41583-019-0132-6.

70. Gao, C., Jiang, J., Tan, Y., and Chen, S. (2023). Microglia in neurodegenerative diseases: mechanism and potential therapeutic targets. Signal Transduct Target Ther 8, 359. 10.1038/s41392-023-01588-0.

71. Blujdea, E.R., van Bokhoven, P., Martino-Adami, P.V., Marshe, V.S., Vromen, E.M., Hok, A.H.Y.S., Boiten, W.A., Irwin, D.J., Chen-Plotkin, A.S., Lemstra, A.W., et al. (2026). Microglia protein profiles in CSF across Alzheimer’s disease clinical stages. Nat Aging 6, 520–533. 10.1038/s43587-026-01088-0.

72. Jorfi, M., Maaser-Hecker, A., and Tanzi, R.E. (2023). The neuroimmune axis of Alzheimer’s disease. Genome Med 15, 6. 10.1186/s13073-023-01155-w.

73. Takata, K., Ginhoux, F., and Shimohama, S. (2021). Roles of microglia in Alzheimer’s disease and impact of new findings on microglial heterogeneity as a target for therapeutic intervention. Biochem Pharmacol 192, 114754. 10.1016/j.bcp.2021.114754.

74. Deczkowska, A., Keren-Shaul, H., Weiner, A., Colonna, M., Schwartz, M., and Amit, I. (2018). Disease-Associated Microglia: A Universal Immune Sensor of Neurodegeneration. Cell 173, 1073–1081. 10.1016/j.cell.2018.05.003.

75. Tsartsalis, S., Sleven, H., Fancy, N., Wessely, F., Smith, A.M., Willumsen, N., Cheung, T.K.D., Rokicki, M.J., Chau, V., Ifie, E., et al. (2024). A single nuclear transcriptomic characterisation of mechanisms responsible for impaired angiogenesis and blood-brain barrier function in Alzheimer’s disease. Nat Commun 15, 2243. 10.1038/s41467-024-46630-z.

76. Eisenmenger, L.B., Peret, A., Famakin, B.M., Spahic, A., Roberts, G.S., Bockholt, J.H., Johnson, K.M., and Paulsen, J.S. (2023). Vascular contributions to Alzheimer’s disease. Transl Res 254, 41–53. 10.1016/j.trsl.2022.12.003.

77. Davoody, S., Asgari Taei, A., Khodabakhsh, P., and Dargahi, L. (2024). mTOR signaling and Alzheimer’s disease: What we know and where we are? CNS Neurosci Ther 30, e14463. 10.1111/cns.14463.

78. Mwale, P.F., Hsieh, C.T., Yen, T.L., Jan, J.S., Taliyan, R., Yang, C.H., and Yang, W.B. (2025). Chitinase-3-like-1: a multifaceted player in neuroinflammation and degenerative pathologies with therapeutic implications. Mol Neurodegener 20, 7. 10.1186/s13024-025-00801-8.

79. Pelkmans, W., Shekari, M., Brugulat-Serrat, A., Sanchez-Benavides, G., Minguillon, C., Fauria, K., Molinuevo, J.L., Grau-Rivera, O., Gonzalez Escalante, A., Kollmorgen, G., et al. (2024). Astrocyte biomarkers GFAP and YKL-40 mediate early Alzheimer’s disease progression. Alzheimers Dement 20, 483–493. 10.1002/alz.13450.

80. van Harten, A.C., Wiste, H.J., Weigand, S.D., Mielke, M.M., Kremers, W.K., Eichenlaub, U., Dyer, R.B., Algeciras-Schimnich, A., Knopman, D.S., Jack, C.R., Jr., and Petersen, R.C. (2022). Detection of Alzheimer’s disease amyloid beta 1-42, p-tau, and t-tau assays. Alzheimers Dement 18, 635–644. 10.1002/alz.12406.

81. Tosun, D., Demir, Z., Veitch, D.P., Weintraub, D., Aisen, P., Jack, C.R., Jr., Jagust, W.J., Petersen, R.C., Saykin, A.J., Shaw, L.M., et al. (2022). Contribution of Alzheimer’s biomarkers and risk factors to cognitive impairment and decline across the Alzheimer’s disease continuum. Alzheimers Dement 18, 1370–1382. 10.1002/alz.12480.

82. Hanseeuw, B.J., Betensky, R.A., Jacobs, H.I.L., Schultz, A.P., Sepulcre, J., Becker, J.A., Cosio, D.M.O., Farrell, M., Quiroz, Y.T., Mormino, E.C., et al. (2019). Association of Amyloid and Tau With Cognition in Preclinical Alzheimer Disease: A Longitudinal Study. JAMA Neurol 76, 915–924. 10.1001/jamaneurol.2019.1424.

83. Coupe, P., Manjon, J.V., Lanuza, E., and Catheline, G. (2019). Lifespan Changes of the Human Brain In Alzheimer’s Disease. Sci Rep 9, 3998. 10.1038/s41598-019-39809-8.

84. Henneman, W.J., Sluimer, J.D., Barnes, J., van der Flier, W.M., Sluimer, I.C., Fox, N.C., Scheltens, P., Vrenken, H., and Barkhof, F. (2009). Hippocampal atrophy rates in Alzheimer disease: added value over whole brain volume measures. Neurology 72, 999–1007. 10.1212/01.wnl.0000344568.09360.31.

85. Qiang, Q., Skudder-Hill, L., Toyota, T., Huang, Z., Wei, W., Adachi, H., and Alzheimer’s Disease Neuroimaging, I. (2024). CSF 14-3-3 zeta(zeta) isoform is associated with tau pathology and cognitive decline in Alzheimer’s disease. J Neurol Sci 457, 122861. 10.1016/j.jns.2023.122861.

86. Soldan, A., Oh, S., Ryu, T., Pettigrew, C., Zhu, Y., Moghekar, A., Xiao, M.F., Pontone, G.M., Albert, M., Na, C.H., and Worley, P. (2023). NPTX2 in Cerebrospinal Fluid Predicts the Progression From Normal Cognition to Mild Cognitive Impairment. Ann Neurol 94, 620–631. 10.1002/ana.26725.

87. Mravinacova, S., Alanko, V., Bergstrom, S., Bridel, C., Pijnenburg, Y., Hagman, G., Kivipelto, M., Teunissen, C., Nilsson, P., Matton, A., and Manberg, A. (2024). CSF protein ratios with enhanced potential to reflect Alzheimer’s disease pathology and neurodegeneration. Mol Neurodegener 19, 15. 10.1186/s13024-024-00705-z.

88. Nilsson, J., Pichet Binette, A., Palmqvist, S., Brum, W.S., Janelidze, S., Ashton, N.J., Spotorno, N., Stomrud, E., Gobom, J., Zetterberg, H., et al. (2024). Cerebrospinal fluid biomarker panel for synaptic dysfunction in a broad spectrum of neurodegenerative diseases. Brain 147, 2414–2427. 10.1093/brain/awae032.

89. Nilsson, J., Cousins, K.A.Q., Gobom, J., Portelius, E., Chen-Plotkin, A., Shaw, L.M., Grossman, M., Irwin, D.J., Trojanowski, J.Q., Zetterberg, H., et al. (2023). Cerebrospinal fluid biomarker panel of synaptic dysfunction in Alzheimer’s disease and other neurodegenerative disorders. Alzheimers Dement 19, 1775–1784. 10.1002/alz.12809.

90. Terry, R.D., Masliah, E., Salmon, D.P., Butters, N., DeTeresa, R., Hill, R., Hansen, L.A., and Katzman, R. (1991). Physical basis of cognitive alterations in Alzheimer’s disease: synapse loss is the major correlate of cognitive impairment. Ann Neurol 30, 572–580. 10.1002/ana.410300410.

91. Zhang, J., and Zhou, Y. (2018). 14-3-3 Proteins in Glutamatergic Synapses. Neural Plast 2018, 8407609. 10.1155/2018/8407609.

92. Hochmair, J., van den Oetelaar, M.C.M., Ravatt, L., Diez, L., Lemmens, L.J.M., Ponce-Lina, R., Sankar, R., Franck, M., Nolte, G., Semenova, E., et al. (2025). Stoichiometric 14-3-3zeta binding promotes phospho-Tau microtubule dissociation and reduces aggregation and condensation. Commun Biol 8, 1139. 10.1038/s42003-025-08548-0.

93. Cornell, B., and Toyo-Oka, K. (2017). 14-3-3 Proteins in Brain Development: Neurogenesis, Neuronal Migration and Neuromorphogenesis. Front Mol Neurosci 10, 318. 10.3389/fnmol.2017.00318.

94. Li, G., Hsu, L.M., Wu, Y., Bozoki, A.C., Shih, Y.I., and Yap, P.T. (2025). Revealing excitation-inhibition imbalance in Alzheimer’s disease using multiscale neural model inversion of resting-state functional MRI. Commun Med (Lond) 5, 17. 10.1038/s43856-025-00736-7.

95. Maestu, F., de Haan, W., Busche, M.A., and DeFelipe, J. (2021). Neuronal excitation/inhibition imbalance: core element of a translational perspective on Alzheimer pathophysiology. Ageing Res Rev 69, 101372. 10.1016/j.arr.2021.101372.

96. Smith, A.M., Davey, K., Tsartsalis, S., Khozoie, C., Fancy, N., Tang, S.S., Liaptsi, E., Weinert, M., McGarry, A., Muirhead, R.C.J., et al. (2022). Diverse human astrocyte and microglial transcriptional responses to Alzheimer’s pathology. Acta Neuropathol 143, 75–91. 10.1007/s00401-021-02372-6.

97. Keren-Shaul, H., Spinrad, A., Weiner, A., Matcovitch-Natan, O., Dvir-Szternfeld, R., Ulland, T.K., David, E., Baruch, K., Lara-Astaiso, D., Toth, B., et al. (2017). A Unique Microglia Type Associated with Restricting Development of Alzheimer’s Disease. Cell 169, 1276–1290 e1217. 10.1016/j.cell.2017.05.018.

98. Montoliu-Gaya, L., Bian, S., Dammer, E.B., Alcolea, D., Sauer, M., Marta-Ariza, M., Ashton, N.J., Belbin, O., Fuchs, J., Watson, C.M., et al. (2025). Proteomic analysis of Down syndrome cerebrospinal fluid compared to late-onset and autosomal dominant Alzheimer s disease. Nat Commun 16, 6003. 10.1038/s41467-025-61054-z.

99. Hawrylycz, M., Kaplan, E.S., Travaglini, K.J., Gabitto, M.I., Miller, J.A., Ng, L., Close, J.L., Hodge, R.D., Long, B., Mollenkopf, T., et al. (2024). SEA-AD is a multimodal cellular atlas and resource for Alzheimer’s disease. Nat Aging 4, 1331–1334. 10.1038/s43587-024-00719-8.

100. Kanehisa, M., and Goto, S. (2000). KEGG: kyoto encyclopedia of genes and genomes. Nucleic Acids Res 28, 27–30. 10.1093/nar/28.1.27.

101. Robin, X., Turck, N., Hainard, A., Tiberti, N., Lisacek, F., Sanchez, J.C., and Muller, M. (2011). pROC: an open-source package for R and S+ to analyze and compare ROC curves. BMC Bioinformatics 12, 77. 10.1186/1471-2105-12-77.

102. Wickham, H. (2016). ggplot2: Elegant Graphics for Data Analysis. Springer-Verlag New York.

103. Kolde, R. (2025). pheatmap: Pretty Heatmaps.

104. Therneau, T.M. (2026). A Package for Survival Analysis in R.

105. Walter, W., Sanchez-Cabo, F., and Ricote, M. (2015). GOplot: an R package for visually combining expression data with functional analysis. Bioinformatics 31, 2912–2914. 10.1093/bioinformatics/btv300.

106. Ritchie, M.E., Phipson, B., Wu, D., Hu, Y., Law, C.W., Shi, W., and Smyth, G.K. (2015). limma powers differential expression analyses for RNA-sequencing and microarray studies. Nucleic Acids Res 43, e47. 10.1093/nar/gkv007.

107. Carlson, M. (2024). Genome wide annotation for Human, primarily based on mapping using Entrez Gene identifiers.

108. Wu, T., Hu, E., Xu, S., Chen, M., Guo, P., Dai, Z., Feng, T., Zhou, L., Tang, W., Zhan, L., et al. (2021). clusterProfiler 4.0: A universal enrichment tool for interpreting omics data. Innovation (Camb) 2, 100141. 10.1016/j.xinn.2021.100141.

109. Jack, C.R., Jr., Albert, M.S., Knopman, D.S., McKhann, G.M., Sperling, R.A., Carrillo, M.C., Thies, B., and Phelps, C.H. (2011). Introduction to the recommendations from the National Institute on Aging-Alzheimer’s Association workgroups on diagnostic guidelines for Alzheimer’s disease. Alzheimers Dement 7, 257–262. 10.1016/j.jalz.2011.03.004.

110. Mirra, S.S., Heyman, A., McKeel, D., Sumi, S.M., Crain, B.J., Brownlee, L.M., Vogel, F.S., Hughes, J.P., van Belle, G., and Berg, L. (1991). The Consortium to Establish a Registry for Alzheimer’s Disease (CERAD). Part II. Standardization of the neuropathologic assessment of Alzheimer’s disease. Neurology 41, 479–486. 10.1212/wnl.41.4.479.

111. Braak, H., and Braak, E. (1991). Neuropathological stageing of Alzheimer-related changes. Acta Neuropathol 82, 239–259. 10.1007/BF00308809.

112. Pang, Z., Lu, Y., Zhou, G., Hui, F., Xu, L., Viau, C., Spigelman, A.F., MacDonald, P.E., Wishart, D.S., Li, S., and Xia, J. (2024). MetaboAnalyst 6.0: towards a unified platform for metabolomics data processing, analysis and interpretation. Nucleic Acids Res 52, W398–W406. 10.1093/nar/gkae253.

113. Gu, Z., Gu, L., Eils, R., Schlesner, M., and Brors, B. (2014). circlize Implements and enhances circular visualization in R. Bioinformatics 30, 2811–2812. 10.1093/bioinformatics/btu393.

114. Allen Institute for Brain Science, U.o.W.A.s.D.R.C., and Kaiser Permanente Washington Health Research Institute. (2022). 10.7303/9618137.

