## Supplemental Information for "Reproducibility-driven discovery and systematic benchmarking reveal a robust cerebrospinal fluid proteomic signature in Alzheimer’s disease"

**Data used in preparation of this article were obtained from the Alzheimer's Disease Neuroimaging Initiative (ADNI) database (adni.loni.usc.edu). As such, the investigators within the ADNI contributed to the design and implementation of ADNI and/or provided data but did not participate in analysis or writing of this report. A complete listing of ADNI investigators can be found at: http://adni.loni.usc.edu/wp-content/uploads/how_to_apply/ADNI_Acknowledgement_List.pdf

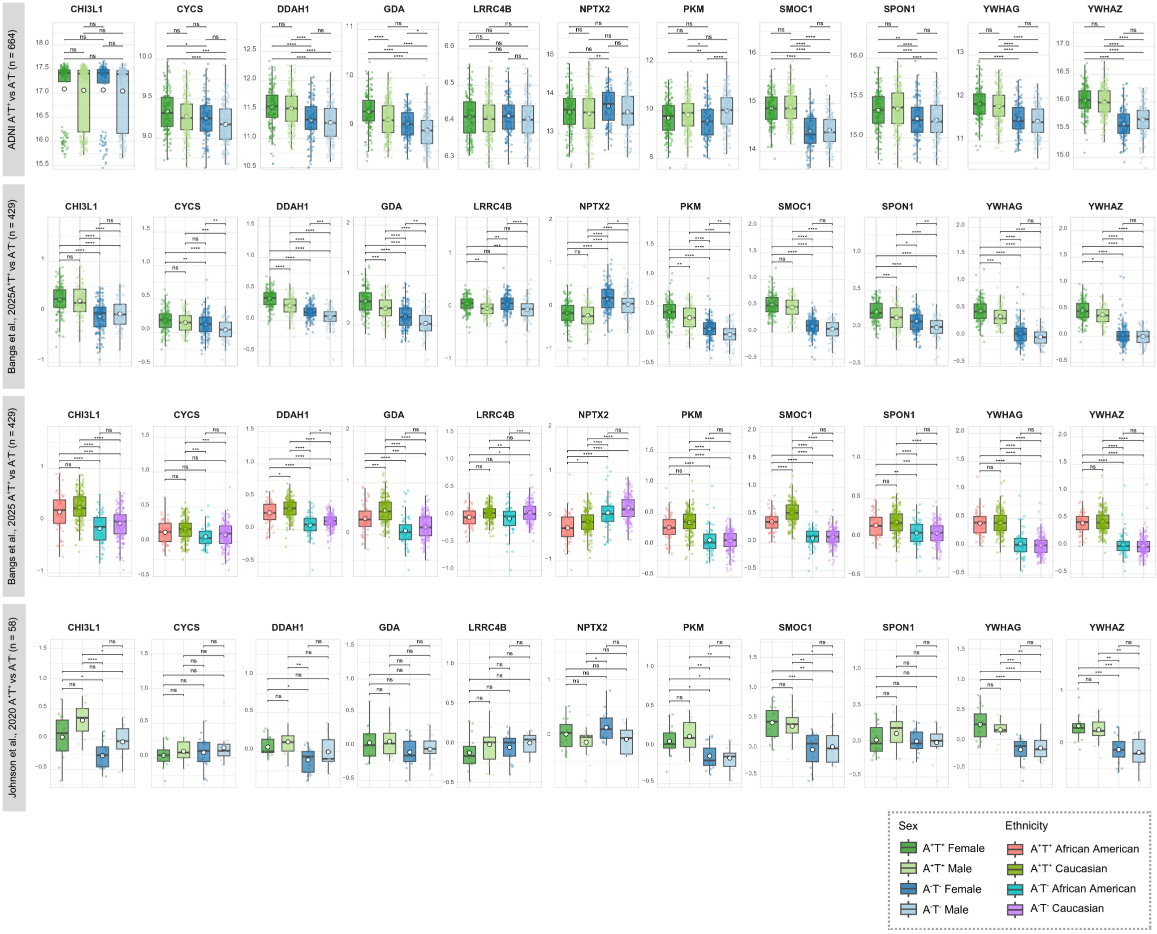


**Figure S1. Differential abundance of PPAV11 proteins between biological Alzheimer's disease (AD) and control subjects stratified by sex and ethnicity.** $Log_2$ abundance of proteins in ADNI (by sex), Bangs et al.[1] (by sex and ethnicity) and Johnson et al.[2] (by sex) comparing $A^+T^+$ and $A^-T^-$ subjects. Box plots represent the median and interquartile range, while mean estimates are by white dots. Significance was assessed by one-way ANOVA followed by Tukey's post hoc test, *p-adjusted < 0.05, **p-adjusted < 0.01, ***p-adjusted < 0.001, ****p-adjusted < 0.0001, ns = non-significant. Abbreviations: ADNI, Alzheimer's Disease Neuroimaging Initiative; A, Amyloid; T, Tau.

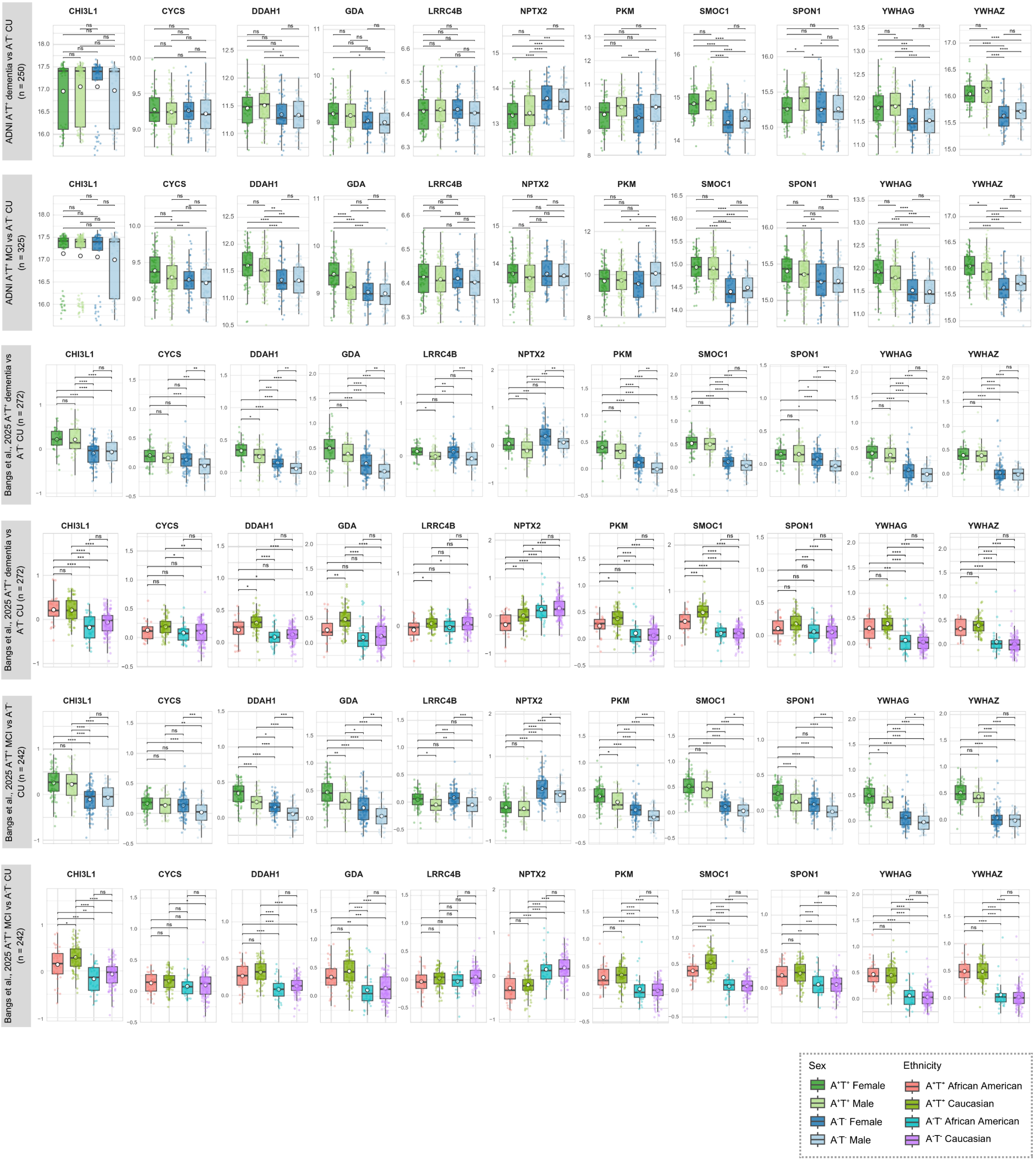


**Figure S2. Differential abundance of PPAV11 proteins between biological and clinical Alzheimer's disease (AD) and control subjects stratified by sex and ethnicity.** $Log_2$ abundance of proteins in ADNI (by sex), Bangs et al.[1] (by sex and ethnicity) and Johnson et al.[2] (by sex) comparing $A^+T^+$ cognitively impaired (MCI or dementia) and $A^-T^-$ CU subjects. Box plots represent the median and interquartile range, while mean estimates are by white dots. Significance was assessed by one-way ANOVA followed by Tukey's post hoc test, *p-adjusted < 0.05, **p-adjusted < 0.01, ***p-adjusted < 0.001, ****p-adjusted < 0.0001, ns = non-significant. Abbreviations: ADNI, Alzheimer's Disease Neuroimaging Initiative; A, Amyloid; T, tau; CU, cognitively unimpaired; MCI, mild cognitively impaired.

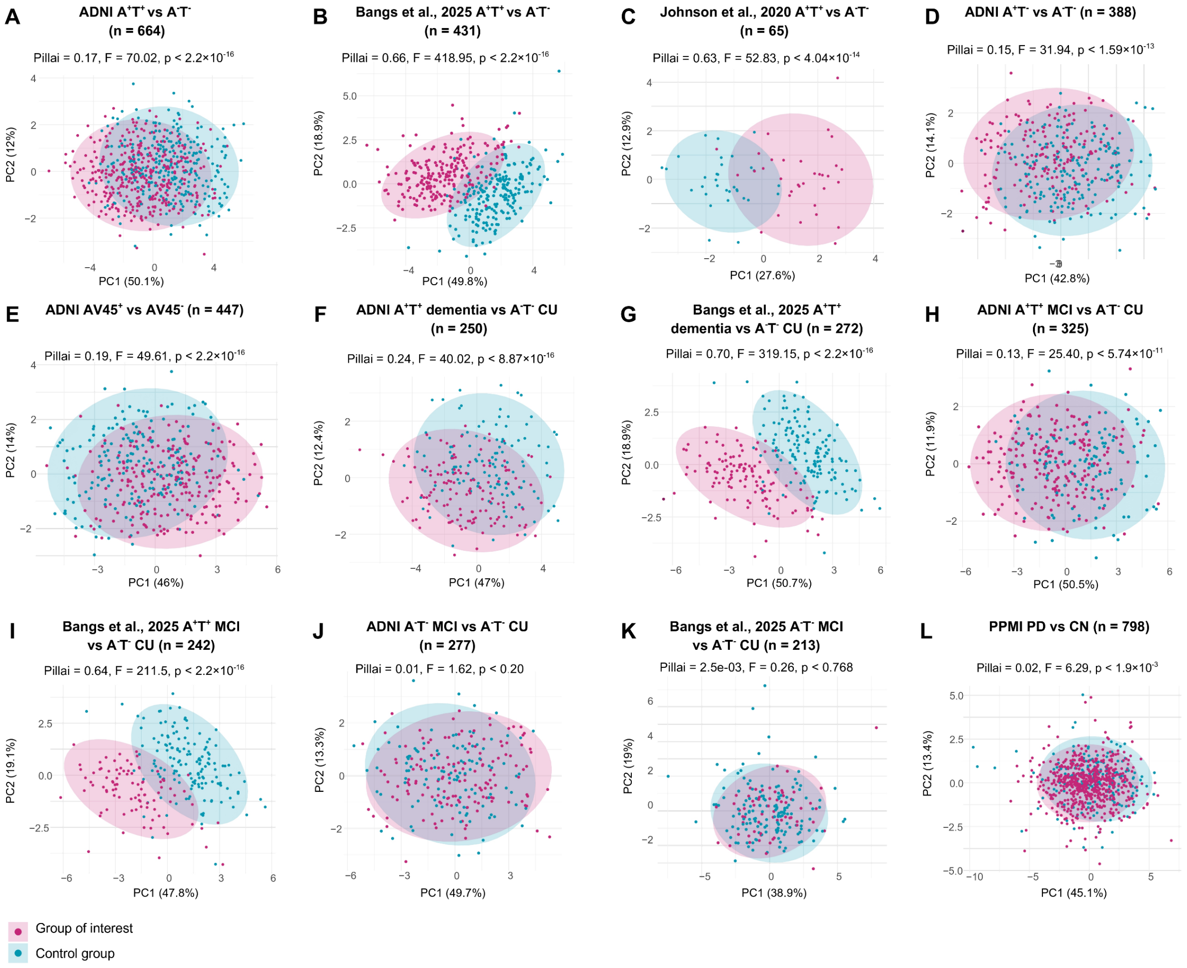


**Figure S3. Differentiation between biological or clinical Alzheimer's disease (AD) and control groups by PPAV11 protein abundances using principal component analysis (PCA).** Subjects stratified by (A-E) biological diagnosis ($A^+T^+$ vs $A^-T^-$, $A^+T^-$ vs $A^-T^-$, and $AV45^+$ vs $AV45^-$); and by (F-I) biological diagnosis and clinical status ($A^+T^+$ MCI vs $A^-T^-$ CU, and $A^+T^+$ dementia vs $A^-T^-$ CU). Selectivity of PPAV11 evaluated by comparing (J-K) $A^-T^-$ MCI vs $A^-T^-$ CU subjects, and (L) PD vs control subjects. Significance was assessed by Pillai's trace (MANOVA). Ellipses represent 95% confidence intervals. Abbreviations: ADNI, Alzheimer's Disease Neuroimaging Initiative; A, Amyloid $\beta_{42}$; T, Tau; AV45, Florbetapir F18 positron emission tomography; MCI, mild cognitively impaired; CU, cognitively unimpaired; PPMI, Parkinson's Progression Markers Initiative; PD, Parkinson's disease; PET, positron emission tomography; CSF, cerebrospinal fluid; MANOVA, multivariate analysis of variance.

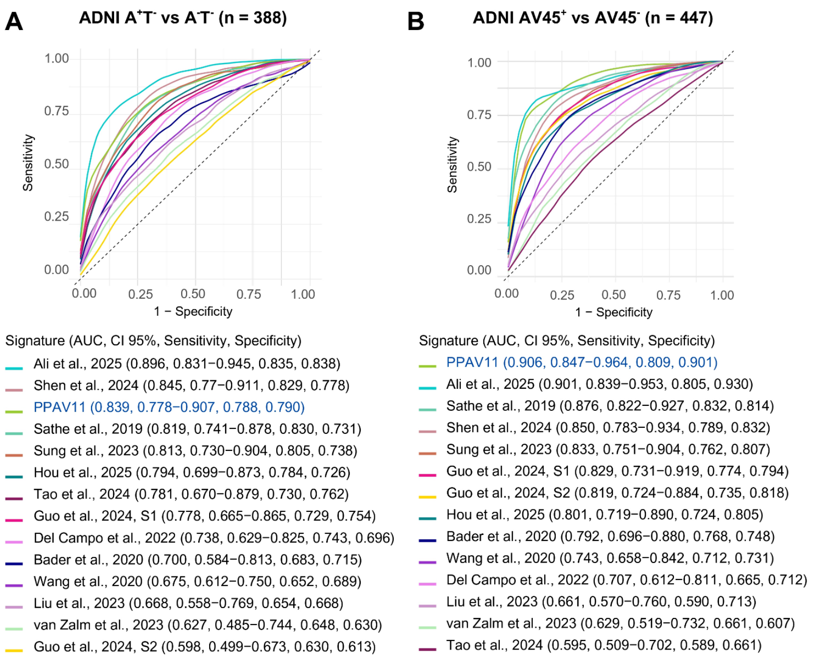


**Figure S4. Receiver operating characteristics (ROC) curve analysis to distinguish between subjects with abnormal and normal brain and CSF levels of Aβ.** Subjects stratified by abnormal and normal in the ADNI cohort by (A) $A\beta_{42}$ CSF levels, and (B) 18F-AV45 brain levels. Comparative analysis was performed against Ali et al.[3], Bader et al.[4], Del Campo et al.[5], Guo et al.[6], Hou et al.[7], Liu et al.[8], Sathe et al.[9], Shen et al.[10], Sung et al.[11], Tao et al.[12], van Zalm et al.[13], and Wang et al.[14]. Sensitivity and specificity were determined by Youden's index. Abbreviations: ADNI, Alzheimer's Disease Neuroimaging Initiative; A, Amyloid $\beta_{42;}$ T, Tau; AV45, Florbetapir F18 positron emission tomography; AUC, area under the curve; CI, confidence intervals; CSF, cerebrospinal fluid.

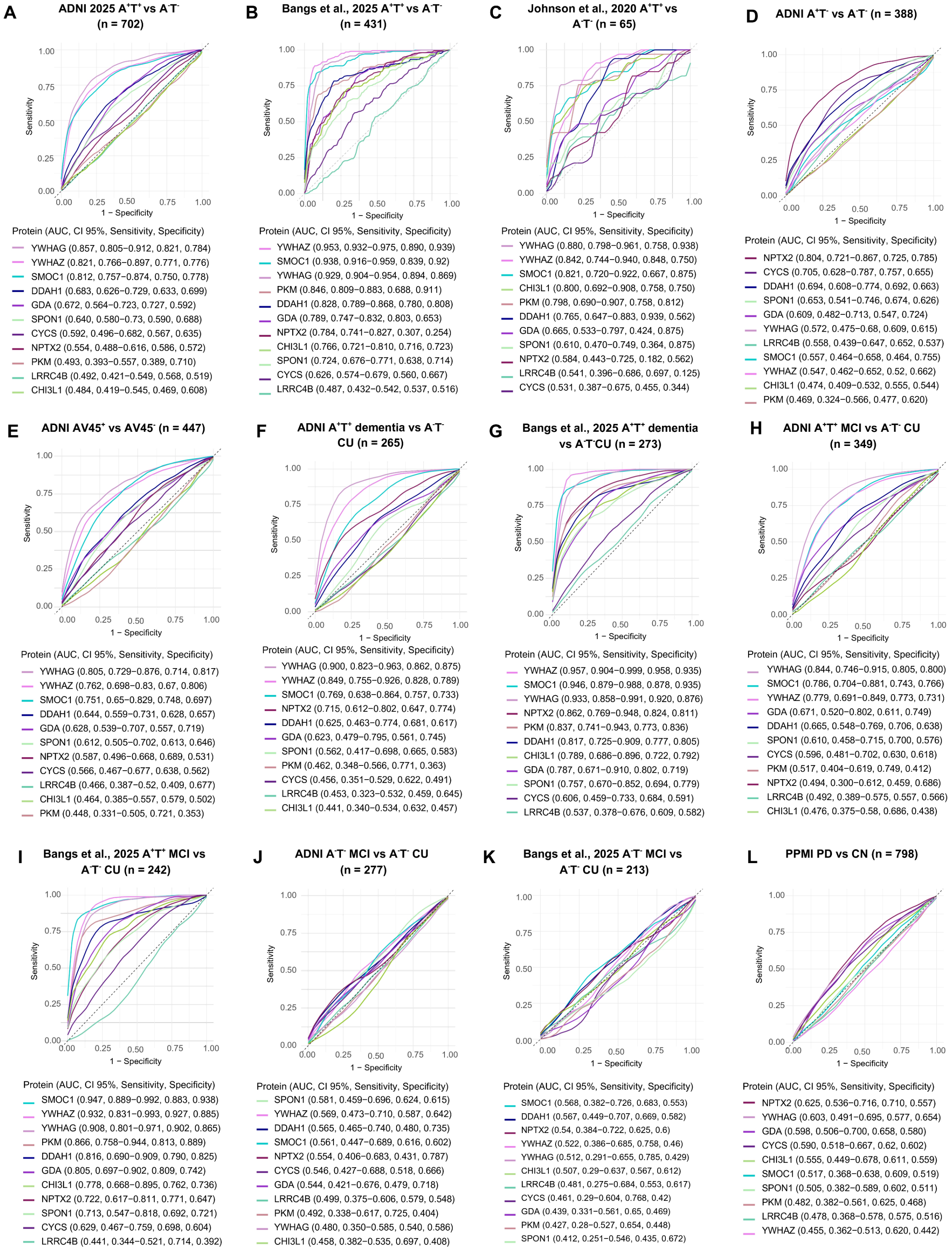


**Figure S5. Validation of individual proteins from PPAV11 signature for biological and clinical Alzheimer's disease (AD) diagnosis**. Receiver operating characteristics (ROC) curve analyses comparing the utility of single proteins to distinguish between subjects stratified by (A-E) biological diagnosis ($A^+T^+$ vs $A^-T^-$, $A^+T^-$ vs $A^-T^-$, and $AV45^+$ vs $AV45^-$); and by (F-I) biological diagnosis and clinical status ($A^+T^+$ MCI vs $A^-T^-$ CU, and $A^+T^+$ dementia vs $A^-T^-$ CU). Selectivity of PPAV11 evaluated by comparing (J-K) $A^-T^-$ MCI vs $A^-T^-$ CU subjects, and (L) PD vs control subjects. Sensitivity and specificity were determined by Youden's index. Abbreviations: ADNI, Alzheimer's Disease Neuroimaging Initiative; A, Amyloid $\beta_{42;}$ T, Tau; AV45, Florbetapir F18 positron emission tomography; CU, cognitively unimpaired; MCI, mild cognitively impaired; PPMI, Parkinson's Progression Markers Initiative; PD, Parkinson's Disease; CN, control; AUC, area under the curve; CI, confidence intervals.

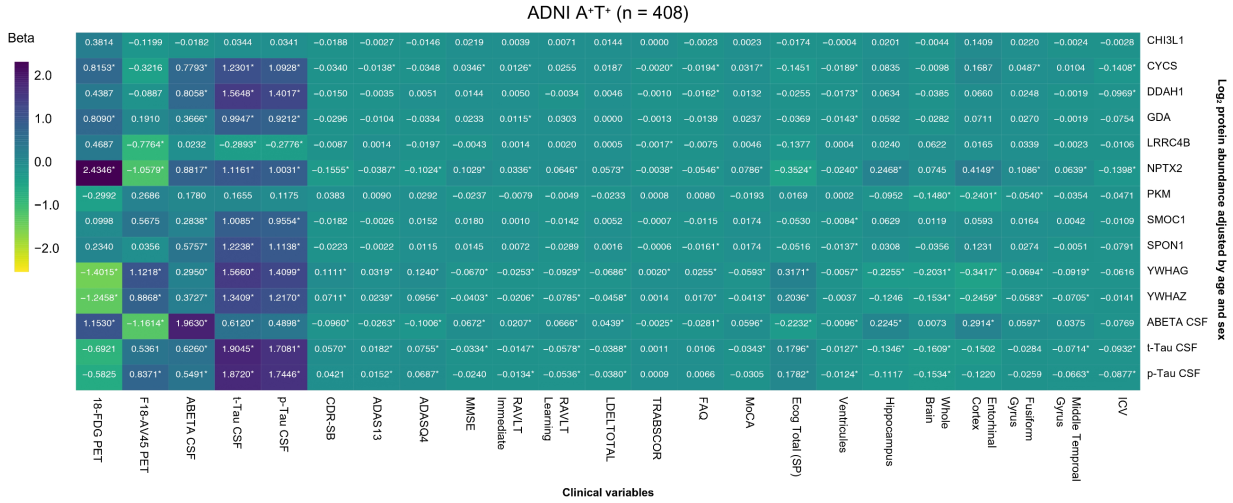


**Figure S6. Correlation analysis of PPAV11 proteins with clinical variables in the ADNI cohort.** Heatmap shows correlations of individual PPAV11 proteins and core AD biomarkers with cognitive tests, brain volume measurements, and biomarkers. Beta coefficient values were derived from LIMMA analysis and represent the association of change in protein abundance (normalized by z-score) per unit of each clinical variable. Models were adjusted for age and sex. MRI volume metrics are expressed in $cm^2$. Significance (p-adjusted < 0.05, Benjamini-Hochberg correction) is highlighted by an asterisk. Abbreviations: ADNI, Alzheimer's Disease Neuroimaging Initiative; 18-FDG PET, $^{18}$F-fluorodeoxyglucose positron emission tomography; F18-AV45 PET, Florbetapir F18 positron emission tomography; t-Tau, total Tau; p-Tau, phosphorylated Tau; CDR-SB, Clinical Dementia Rating–Sum of Boxes; ADAS, Alzheimer's Disease Assessment Scale; MMSE, Mini Mental State Examination; RAVLT, Rey Auditory Verbal Learning Test; LDEDLTOTAL, logical memory delayed recall total; TRABSCOR, Trail Making Test; FAQ, Functional Activities Questionnaire; MoCA, Montreal Cognitive Assessment; ECOG (SP), Everyday Cognition, spouse report; ICV, intracerebroventricular.
